# Maladaptive avoidance learning in the orbitofrontal cortex in adolescents with major depression

**DOI:** 10.1101/2021.05.21.21257570

**Authors:** David Willinger, Iliana I. Karipidis, Selina Neuer, Sophie Emery, Carolina Rauch, Isabelle Häberling, Gregor Berger, Susanne Walitza, Silvia Brem

## Abstract

**Background:** Understanding the mechanisms in the brain’s incentive network that give rise to symptoms of major depressive disorder (MDD) during adolescence provides new perspectives to address MDD in early stages of development. This functional magnetic resonance imaging study determines whether instrumental vigor and brain responses to appetitive and aversive monetary incentives are altered in adolescent MDD and associated with symptom severity.

**Methods:** Adolescents with moderate to severe MDD (n=30, age=16.1 [1.4]), and healthy controls (n=33, age=16.2 [1.9]) matched for age, sex, and IQ performed a monetary incentive delay task. During outcome presentation, prediction error signals were used to study the response and coupling of the incentive network during learning of cue-outcome associations. A computational reinforcement model was used to assess adaptation of response vigor. Brain responses and effective connectivity to model-derived prediction errors were assessed and related to depression severity and anhedonia levels.

**Results:** Participants with MDD behaved according to a more simplistic learning model and exhibited slower learning. Effective connectivity analysis of fMRI data revealed that impaired loss error processing in the orbitofrontal cortex was associated with aberrant gain-control. Anhedonia scores correlated with loss-related error signals in the posterior insula and habenula.

**Conclusions:** Adolescent MDD is selectively related to impaired processing of error signals during loss, but not reward, in the orbitofrontal cortex. Aberrant evaluation of loss outcomes might reflect an early mechanism of how negative bias and helplessness manifest in the brain. This approach sheds light on pathomechanisms in MDD and may improve early diagnosis and treatment selection.

## 1 Introduction

Major depressive disorder (MDD) is among the most prevalent mental health problems in adolescents worldwide(1) with an estimated 12 months prevalence of 7.5% in mid to late adolescence(2). Adolescent MDD increases the risk for substance misuse, can severely impair success in school, social life, and cognitive functions(3), and is a major risk factor for suicide, which is among the leading causes of death at this age(4). Despite these adverse outcomes, relatively little is known about brain mechanisms related to MDD with early onset. Recent evidence suggest that disrupted prediction error (PE) signaling constitutes a potential brain mechanism that promotes the persistence of negative beliefs and anhedonia(5).

It is widely established that the dopaminergic system is fundamental in encoding reward and loss PEs(6), which are crucial in reinforcement learning and decision making. Influential computational models(7, 8) suggest that during the anticipation of an incentive an expected value (Q) signal is generated, which is the product of a learned probability and the magnitude of the incentive. During outcome receipt, the difference between the expected value signal and the actual outcome is signaled as PE to update predictions. While the ventral striatum primarily encodes PEs in reward contexts, the anterior insula does so in avoidance or loss contexts(9). In addition, brain regions sensitive to errors in reward and loss contexts are the orbitofrontal cortex (OFC) and the dorsal anterior cingulate cortex (dACC)(10, 11).

Previous studies have demonstrated that striatal PE signaling is reduced in adult MDD in reward(12, 13) and loss contexts(6). Therefore, it is crucial to establish whether this aberrant signaling is already present in early onset MDD or whether deviations in reward and loss processing are a downstream effect of chronicity and burden(6). Studies in young cohorts suggest that blunted reward sensitivity in the ventral striatum predicts symptom deterioration(14, 15) and is present in individuals at high familial risk for depression(16, 17). In addition, there is emerging evidence of impaired loss sensitivity in the incentive network in high risk groups(18) that predicts future depressive symptoms(19). However, it remains unclear whether atypical learning signals are linked to deficient motivated behavior. Previous studies showed mixed results when applying computational models to behavioral data in adult MDD, with learning rates depending on the task used and the specific learning process probed(20). This clearly indicates that more work is necessary to identify brain mechanisms that give rise to aberrant incentive processing in depression, particularly during development. In this functional magnetic resonance imaging (fMRI) study, we hypothesized that the encoding of reinforcement learning signals is impaired in adolescent MDD. We employed a monetary incentive delay task(10, 21) with varying magnitude (low, high) and valence (reward, loss) to probe the neural circuits supporting PE and expected value processing. On a neural level, we hypothesized (a) decreased reward PE signaling within the striatum in MDD(5, 15), (b) a negative association between anhedonia scores and blunted responses to rewards in the striatum and the OFC(22), and (c) reduced reactivity of the OFC during loss events(19). Behaviorally, we tested whether response vigor, i.e. adaptive responses to reward and losses of varying magnitudes(23), was differentially modulated in MDD, and whether there are differences in the update of value representations in the instrumental learning task.

## 2 Methods and materials

### 2.1 Participants

Thirty adolescent patients and 33 healthy individuals matched for age, IQ, gender, and handedness participated in this study(Table 1). Participants with MDD were recruited through clinical services. All participants underwent a semistructured clinical interview (K-SADS-PL(24) or MINI-KID(25)). Participants with MDD fulfilled a diagnosis according to the DSM-IV (codes 296.20-296.23, 296.30-296.33). Past and present comorbid diagnoses in patients comprised anxiety disorders (n=7), obsessive-compulsive disorder (n=1), and attention-deficit hyperactivity disorder (n=2). Moreover, we assessed a battery of self-report questionnaires, IQ and working memory of all participants (Table 1). We included total scores and scores from the anhedonia subscale from the German version of the Child Depression Inventory(26) (CDI) in our neuroimaging analyses. Healthy controls (HC) were recruited through schools and volunteer websites. For controls, exclusion criteria comprised any current psychiatric disorder, other major medical illnesses, drug abuse, any MRI contraindication, pregnancy, and a history of brain injury. Three control participants had a past diagnostic work-up for ADHD but they were currently symptom-free and were not taking any medication during the study. All procedures contributing to this work comply with the ethical standards of the ethic committee of the Canton of Zurich, Switzerland (BASEC 2017-02179) and with the Helsinki Declaration of 1975, as revised in 2008. All participants gave their written informed consent, parents or legal guardians gave signed informed consent for children under the age of 14 years. They were reimbursed for participation and informed about the opportunity to additionally win up to CHF 20 during the task.

**Table 1.** Clinical and demographic characteristics of study participants.

|  | **Controls** | **MDD** | **Test statistic** | **p value<sup>a</sup>** |
| --- | --- | --- | --- | --- |
| **Age (years), range (min-max)** | 16.2 (1.9),<br>11.2-18.8 | 16.1 (1.4),<br>12.8-18.7 | $U=553.5$ | .425 |
| **Sex (males), No. (%)** | 10 (30%) | 10 (33%) | $\chi^2(1)=0.07$ | .796 |
| **Handedness (right), No. (%)** | 32 (97%) | 28 (93%) | $\chi^2(1)=0.46$ | .500 |
| **In-scanner movement (FD, mm)** | 0.18 (0.09) | 0.18 (0.08) | $t(61)=0.09$ | .930 |
| **CD-RISC** | 72.9 (10.1) | 38.6 (15.6) | $t(58)=10.16$ | <.001 |
| **CDI** | 8.4 (6.6) | 29.6 (9.3) | $U=38.0$ | <.001 |
| **Anhedonia** | 2.3 (2.2) | 10.5 (2.8) | $U=13.5$ | <.001 |
| **Negative mood** | 2.2 (2.0) | 6.4 (2.4) | $U=88.0$ | <.001 |
| **Negative self-esteem** | 1.0 (1.2) | 5.0 (1.7) | $U=42.0$ | <.001 |
| **Ineffectiveness** | 1.2 (1.2) | 5.0 (1.9) | $U=54.5$ | <.001 |
| **Interpersonal problems** | 1.1 (1.2) | 3.7 (1.5) | $U=74.5$ | <.001 |
| **Stomach** | 0.6 (0.6) | 1.1 (0.8) | $U=301.5$ | .018 |
| **RIAS IQ** | 104.5 (6.9) | 108.0 (8.7) | $t(60)=-1.75$ | .079 |
| **PSS** | 22.4 (6.6) | 28.8 (7.7) | $t(57)=-3.44$ | .001 |
| **SDQ** | 8.8 (5.3) | 16.3 (5.6) | $t(56)=-5.26$ | <.001 |
| **WISC-IV Digitspan (forward)** | 8.9 (2.1) | 8.8 (2.0) | $t(60)=0.32$ | .747 |
| **WISC-IV Digitspan (backward)** | 8.6 (1.6) | 9.4 (2.0) | $t(60)=-1.70$ | .094 |
| **WISC-IV Mosaic,** | 57.0 (5.7) | 59.0 (6.2) | $t(56)=-1.27$ | .208 |
| **Current Medication, No. (%)** |  |  |  |  |
| **No medication** | NA | 10 (33%) | NA | NA |
| **SSRI** | NA | 18 (60%) | NA | NA |
| **Dual-action antidepressant<sup>b</sup>** | NA | 2 (7%) | NA | NA |
| **NERI** | NA | 2 (7%) | NA | NA |
| **Antipsychotic<sup>c</sup>** | NA | 2 (7%) | NA | NA |
| **Methylphenidate** | NA | 2 (7%) | NA | NA |

Data are presented as mean (SD) if not indicated otherwise.
Abbreviations: CD-RISC, Connor-Davidson Resilience Scale; CDI, Children Depression Inventory; FD, framewise displacement; RIAS, Reynolds Intellectual Assessment Scales; PSS, Perceived Stress Scale; SDQ-K, Strength and Difficulty Questionnaire for Children; WISC, Wechsler Intelligence Scale for Children.
<sup>a</sup>Uncorrected *p* values for between-group comparisons; significance threshold *p*<.05.
<sup>b</sup>Serotonin-noradrenalin reuptake inhibitor
<sup>c</sup>Used for behavioral control

### 2.2 Experimental task

In this study, we used the Monetary Incentive Delay task (MID, Figure S1) task to examine incentive processing in different valence and magnitude contexts(21). This fMRI task minimizes cognitive confounds (e.g. solving a task with different strategies) due to the simple decision processes(27). Importantly, it allowed us to assess individual trial-by-trial reward and loss prediction errors based on the associations participants built up relying on the outcome of the preceding trials, allowing us to distinguish it from outcome processing. Every trial began with the presentation of a cue indicating the level of magnitude (CHF 1, CHF 4) and the valence (reward, loss-avoidance, null) for a button press on time (“hit”). Participants were instructed to use the index finger of their dominant hand to press a button on a two-button fibre-optic response pad (Current Design Inc., Philadelphia, PA) as soon as the go-signal target symbol, a star, appeared. In total, each cue was presented 24 times (i.e. 120 trials in total, mean stimulus onset asynchrony = 8500ms, 7750 – 10750*ms*), in two separate MRI runs. We used an adaptive algorithm that adjusted the presentation times of the target to the response time of the participant to ensure a hit rate of ∼66%. The cue symbols indicating valence (square, triangle, and circle) were counterbalanced across subjects. A filled symbol indicated a trial with high magnitude (CHF 4), whereas an empty symbol indicated trials with low magnitude (CHF 1). All participants had a short training session outside the scanner (∼2 minutes) to familiarize themselves with the task and we ensured that cue-outcome contingencies were understood. The task was implemented in python (pygame, https://www.pygame.org) and presented using video goggles (VisuaStimDigital, Resonance Technology, Northridge, CA) with a resolution of 800×600px.

Post-scan ratings of subjective liking and arousal for rewards and losses were assessed at the end of the scanning session. Participants rated their subjective liking (“How much did you like this outcome?”) and arousal (“How excited were you by the outcome?”) on a continuous scale using a slider between 0 (strongly dislike, not aroused) and 100 (strongly like, highly aroused).

### 2.3 Computational Modeling

The task employed here allows to assess mechanisms that determine behavior (i.e. instrumental response vigor). To assess these quantities we constructed several competing generative behavioral models that predicted trial-by-trial reaction times for each participant. This allowed us to identify parameters with mechanistic meaning for observed response vigor, the latent representation of value, and the participant-specific learning rate. Even though no learning is necessary to perform this task well, previous studies have found that learning signals continue to be generated even if an association is well-learned (28).

The winning model included the parameters *cue salience*, i.e. the magnitude of possible outcomes × probability, *novelty*, i.e. the unsigned PE, a *post-error* term, a *linear* term, and the intercept. Group differences between controls and patients were assessed for parameters of the best-fitting model across both groups. A detailed description of the behavioral model, model simulations and the behavioral analysis is provided in the Supplement (Figure S2-3, Table S1-4).

### 2.4 Functional MRI analysis

#### 2.4.1 Model-based fMRI - GLM analysis

To investigate the trial-by-trial effect of the computational variables derived from the computational model, we used the variables of the best behavioral model across participants (Table S2) and entered expected value and prediction error as parametric modulators for cue onset and feedback, respectively (Supplemental methods). Group effects were assessed with two-sample *t*-tests, where we entered the contrast images for the expected values and the prediction errors for healthy controls and MDD patients. The cluster-level significance threshold for the whole-brain group analyses was set to p_FWEc_ < 0.05 with a cluster-defining threshold of p_CDT_<0.001. All fMRI analyses were conducted in SPM12 (version 7487), labels for brain regions are based on the Automated Anatomical Atlas (29).

#### 2.4.2 Effective connectivity analysis

To reveal the functional coupling between regions of the incentive network in participants with and without MDD, we performed a dynamic causal modeling (DCM) analysis(30, 31). The regions for this analysis were selected based on (a) previously published findings of incentive processing(9, 10, 32), (b) findings of studies in participants with a history and at risk for MDD(18, 19) and (c) our second-level general linear model (GLM) analyses (Figure **1**A, Table S5-8, Figure S4-6). The aforementioned studies have provided compelling evidence that the insula and the dACC play a significant role in loss-avoidance learning, a finding we corroborate across groups during reward and loss processing. Furthermore, we found a significant group effect in the OFC, that suggested aberrant network dynamics in MDD.

**Figure 1.**
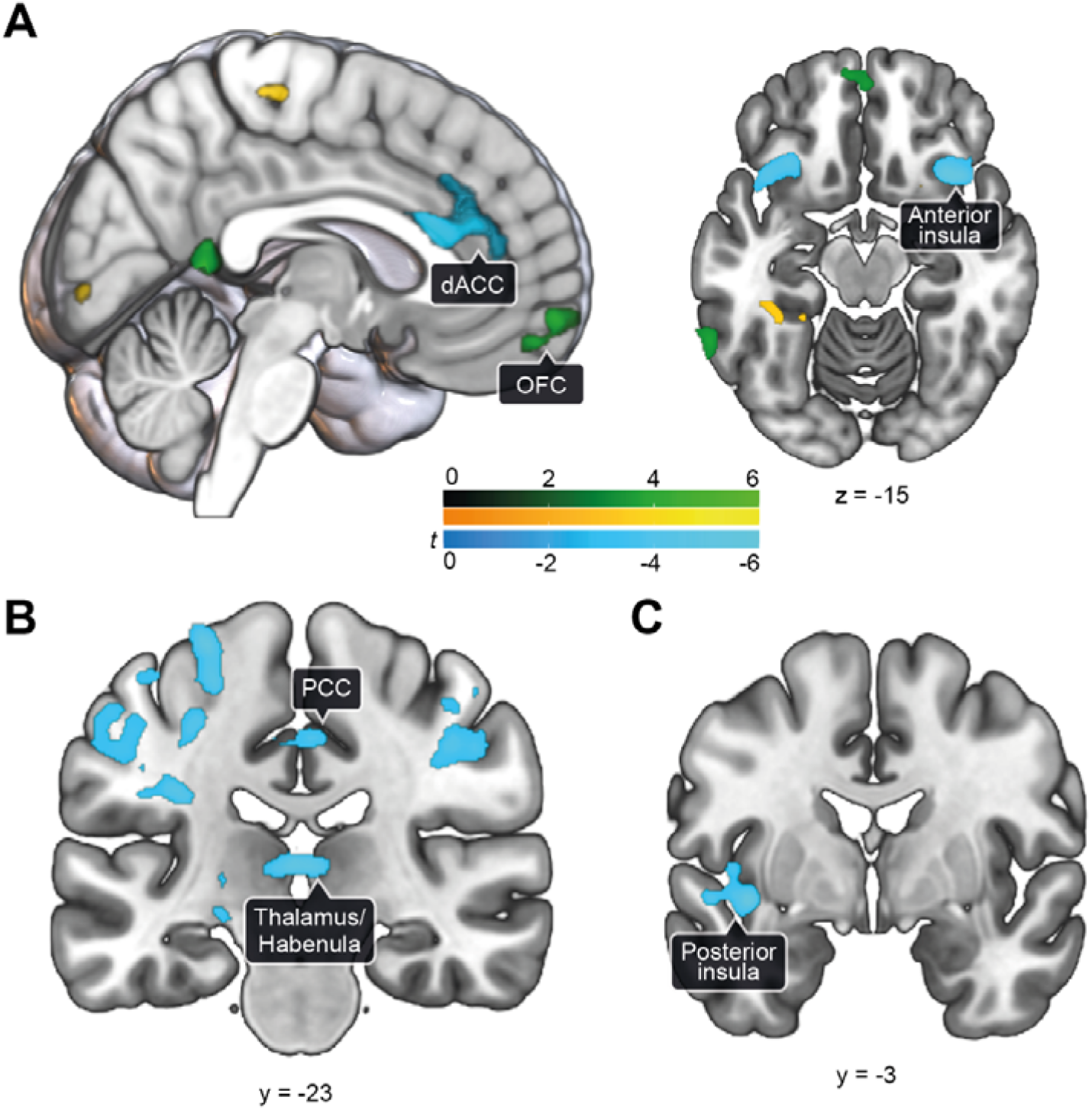
(A) Effect of loss outcome error 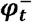 during feedback presentation across all participants (*N*=63). Patients showed significantly reduced responses related to errors during loss compared to controls in the orbitofrontal cortex (OFC, green). Consistent effects across groups (positive effect in yellow, negative effect in blue) of outcome error were found in the dorsal anterior cingulate cortex (dACC) and the anterior insula. Details are reported in Table 2 and main effects in Table S5. *p*_FWEc_ <.05, *p*_CDT_<.001. **(B) Assessment of the effect of anhedonia within patients (*n*=29)**. A negative relationship between magnitude-modulated loss prediction error 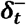-related actitivity and anhedonia scores was observed in a cluster in the medial thalamus / habenula and the posterior cingulate cortex (PCC). **(C)** A significant negative association between anhedonia scores and loss outcome error 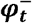 related activity was found in the posterior insula.

For the DCM analysis, we localized the effects of hits and misses across trials, by performing whole-brain contrasts with the CDI as covariate (Figure S5) and extracted the timeseries for each subject from activated voxels (*p*<0.05, uncorrected) within a 12mm spherical search volume around the group maxima from the miss-hit (insula [x=35, y=18, z=-18mm MNI]; dACC [x=7, y=36, z =30mm MNI], Table S8) and the all-events (IOG [x=29; y=-80, z=-16mm MNI], Table S10) contrast. The search volume for the OFC comprised the voxels in the active OFC [x=-7;y=46;z=-16mm MNI] cluster of the second-level hit-miss contrast (Table S8, cluster-extent threshold *p*_FWEc_ < .05). If a participant’s maximum within the search volume differed from the group maximum, we centered the sphere around the participant’s maximum. The first eigenvariate of the time course of all active voxels (*p* < 0.05) was then extracted and adjusted for any motion effects. One patient and two controls had to be excluded from this analysis, as they did not show any activation in the dACC for the defined threshold.

The feedback regressor was the driving input for the visual region. The model comprised direct forward connections from the visual area to all other fully interconnected regions. Since the GLM analysis revealed loss-related group differences in OFC activity (see Results), our main interest was to study network effects during the loss condition, nevertheless, the reward condition was also included to improve the model fit. We included contextual modulation of prediction errors, magnitude (−1 for low and +1 for high), and their interaction term, i.e. magnitude–sensitive PEs on the self-connections of the regions (see Supplement). In this model, the self-connections embody the change in synaptic gain for a given task context(31). Here, our goal was to identify the network dynamics that give rise to the lower error signal in the OFC in MDD patients during loss processing. For this, we set up a DCM analysis within the Parametric Empirical Bayes (PEB) framework.

On the first level, the full DCM model of each participant was estimated iteratively in an empirical Bayesian inversion scheme(31). The individual DCM parameters from the first-level were then entered in the second-level PEB model to determine (a) the differences between the MDD group and controls and (b) the group mean. This analysis was carried out separately for intrinsic and modulatory connections. We performed Bayesian model reduction to iteratively discard those model parameters not contributing to the model evidence. Then, we averaged the parameters of the best PEB models weighted by the posterior probability of the respective model. A PEB model parameter was considered significant when exceeding a 95% posterior probability of being present vs absent based on the model evidence(31, 33). Leave-one-out cross-validation was used to assess whether the predicted and actual group effect showed an independent out-of-sample correlation.

## 3 Results

### 3.1 Altered learning of cue-outcome associations in MDD

Bayesian model comparison revealed that the response model including *cue salience* and *novelty* terms using a single learning rate fitted the response data best across groups. Nevertheless, we found that in patients the simpler model with a single learning rate performed better (posterior probability, PP=57.7%, Table S2), whereas a more complex model with separate learning rates for rewards and losses fitted data better in controls only (PP=51.6%). However, while we found this difference among groups, the difference between the simple and the more complex model was not very strong (Bayes factor = 2.00) and thus we decided to continue the analysis with the simpler model, as this performed best for both groups. A between-group comparison of the learning rate of the best-fitting model across all participants showed that the learning rate was marginally lower in MDD (α:MDD,0.050 [0.021]; controls, 0.052 [0.012]; U=617; *p*=.095). This difference was significant after removing one outlier from to the MDD group (Grubb’s test: *G*=6.107, *U*=0.389, *p*<10^−11^; α:MDD, .046□[.009]; controls, .052□[.012]; t(60)=2.04; *p*=.046, Supplementary Methods). The response model parameters, in particular *cue salience* and *novelty*, did not differ between groups (all *p*>.10, Table S4). These results demonstrate a non-discriminable value update mechanism underlying adolescent MDD for both reward and loss conditions, while there was positive evidence that controls’ response vigor was best described by a more flexible dual update model for both valences. In addition, the comparison of the *learning rate* shows that MDD participants changed their value expectations slower across the task.

### 3.2 OFC gain control explains atypical aversive outcome signaling in MDD

When processing loss feedback, participants with MDD showed a significantly lower response to the outcome error signal 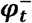 in the OFC (Figure **1**, Table 2). While this region reflected a signal encoding the difference between subjective belief of the outcome and the actual outcome in reward and loss in controls (Table S5), patients only expressed this signal during rewarding and not loss-avoidance trials.

**Table 2.** Group differences between MDD patients and healthy controls for 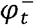 processing

| Brain region | MNI coordinates |  |  | Significant activation |  | Peak |
| --- | --- | --- | --- | --- | --- | --- |
| | x | y | z | $p_{\text{FWEc}}$ | k | Z |
| **Controls > MDD** |  |  |  |  |  |  |
| Precuneus_L | -3 | -46 | 10 | .022 | 128 | 4.67 |
| Frontal_Sup_2_R | 17 | 28 | 44 | .046 | 108 | 4.48 |
| Temporal_Mid_L | -67 | -50 | -6 | .000 | 269 | 4.24 |
| Angular_L | -55 | -64 | 24 | .006 | 166 | 4.08 |
| Temporal_Mid_R | 65 | -10 | -16 | .034 | 116 | 4.05 |
| Frontal_Med_Orb_L | -3 | 60 | -8 | .019 | 132 | 3.93 |
| **MDD > Controls** |  |  |  |  |  |  |
| NS |  |  |  |  |  |  |

Abbreviations: k, cluster size; R, right; L, left; MNI, Montreal Neurological Institute.

To further scrutinize the origin of this effect, we performed a DCM analysis (Figure 2, Table 3). Bayesian model averaging showed that the effect of loss-magnitude on the self-inhibition parameter of the OFC was significantly more negative in MDD, i.e. the region was more disinhibited during processing the outcome of high compared to low loss. These results indicate that MDD is related to aberrant gain control in the OFC, specifically in high loss contexts. Moreover, the self-inhibition of the dACC was significantly lower across task conditions in the MDD group. The posterior mean of the group effect on the self-inhibition of the OFC was significantly related to the learning rate across all participants (Spearman’s *ρ*=.295, *p*=.023, *n*=59), but not in the dACC (*ρ*=-.135, *p*=.301, *n*=59). A leave-one-out cross-validation using the loss-magnitude dependent difference in self-connection strength in the OFC explained a significant amount of the inter-subject variability between MDD and controls, showing an independent out-of-sample correlation of *r(*58)=0.38, *p*=0.001.

**Table 3.** Average connectivity during feedback phase obtained by Bayesian model averaging of PEB model parameters

| Connection type | Commonalities | PP<br>Commonalities | Depression | PP<br>Depression |
| --- | --- | --- | --- | --- |
| **Endogenous parameters** |  |  |  |  |
| OFC→Insula | 0 | 0 | 0 | 0 |
| OFC→dACC | -0.195 | 1* | 0.04 | 0.66 |
| dACC→Insula | 0.812 | 1* | 0 | 0 |
| dACC→OFC | -0.345 | 1* | 0 | 0 |
| Insula→OFC | 0.284 | 1* | 0 | 0 |
| Insula→dACC | -0.084 | 1* | -0.049 | 0.85 |
| IOG→Insula | 0.144 | 1* | 0 | 0 |
| IOG→dACC | -0.228 | 1* | 0.017 | 0.53 |
| IOG→OFC | -0.108 | 1* | -0.013 | 0.58 |
| **Self-inhibition parameters** |  |  |  |  |
| OFC→OFC | -0.467 | 1* |  |  |
| Insula→Insula | 0.311 | 1* |  |  |
| dACC→dACC | -0.288 | 1* | -0.139 | 1* |
| IOG→IOG | 1.740 | 1* |  |  |
| **Modulatory parameters** |  |  |  |  |
| Insula→Insula, $\varphi_t^+$ | 0 | 0 | 0 | 0 |
| Insula→Insula, $M^+$ | 0 | 0 | 0 | 0 |
| Insula→Insula, $\varphi_t^+ \times M^+$ | 0 | 0 | 0 | 0 |
| Insula→Insula, $\varphi_t^-$ | 0 | 0 | 0 | 0 |
| Insula→Insula, $M^-$ | 0 | 0 | 0 | 0 |
| Insula→Insula, $\varphi_t^- \times M^-$ | -0.482 | 1* | 0 | 0 |
| dACC→dACC, $\varphi_t^+$ | 1.506 | 1* | 0 | 0 |
| dACC→dACC, $M^+$ | -0.103 | 0.55 | 0 | 0 |
| dACC→dACC, $\varphi_t^+ \times M^+$ | 0 | 0 | 0.194 | 0.89 |
| dACC→dACC, $\varphi_t^-$ | 1.986 | 1* | 0 | 0 |
| dACC→dACC, $M^-$ | -0.628 | 1* | 0 | 0 |
| dACC→dACC, $\varphi_t^- \times M^-$ | -0.308 | 0.99* | 0 | 0 |
| OFC→OFC, $\varphi_t^+$ | 1.506 | 1* | 0.351 | 0.90 |
| OFC→OFC, $M^+$ | 0.148 | 0.54 | 0 | 0 |
| OFC→OFC, $\varphi_t^+ \times M^+$ | 0 | 0 | -0.140 | 0.51 |
| OFC→OFC, $\varphi_t^-$ | -0.748 | 1* | 0 | 0 |
| OFC→OFC, $M^-$ | -0.495 | 1* | -0.663 | 1* |
| OFC→OFC, $\varphi_t^- \times M^-$ | 0.781 | 1* | 0 | 0 |
| **Input parameter** |  |  |  |  |
| Feedback→IOG | 4.287 | 1* | -0.208 | 0.64 |

Between-region connections are in units of Hz. Self-connections, where the source and target are the same, are the log of scaling parameters that multiply up or down the default value $-0.5\text{Hz}$ . $n = 60$ . dACC, dorsal anterior cingulate cortex; INS, insula; IOG, inferior occipital gyrus; LPFC, lateral prefrontal cortex; M, magnitude; $\varphi$ , error signal; THL, thalamus; VS, ventral striatum; PP, posterior probability.

**Figure 2.**
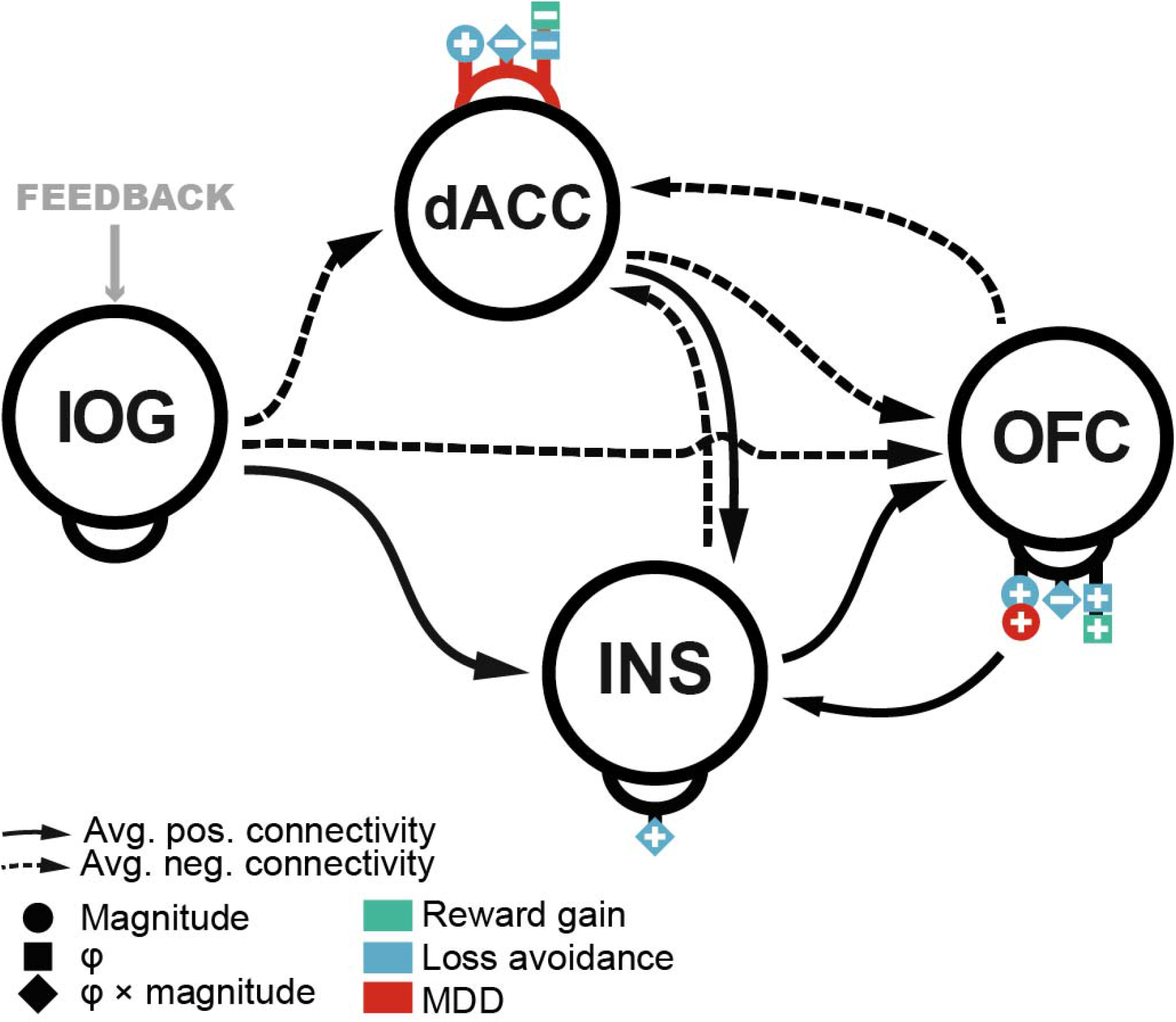
Effective connectivity during reward and loss feedback processing. There was a significant group effect of the factor *loss magnitude* on the self-inhibition parameter in the OFC, indicating aberrant input sensitivity during feedback in loss avoidance contexts in adolescent MDD. Cross-validation showed that this effective connectivity parameter was able to predict the group variable indicated by a significant out-of-sample correlation. Importantly, this parameter was associated with the learning rate of participants, which was lower in adolescents with MDD. The arrows reflect the posterior estimates of the second level PEB model after Bayesian model reduction. Self-connections are depicted as half-circle on each region. Solid lines indicate positive effective connectivity whereas dashed lines represent negative effective connectivity. Details of the results are reported in Table 3. Abbreviations: IOG, inferior occipital gyrus; dACC, dorsal anterior cingulate cortex; INS, anterior insula; MDD, major depressive disorder; OFC, orbitofrontal cortex; φ, outcome error.

### 3.3 Neural correlates of depression severity and anhedonia

Regression analysis of anhedonia scores within patients revealed significant associations in brain signaling of learning variables for loss. Particularly, we found that magnitude-modulated loss prediction error signaling 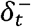 was associated negatively with anhedonia scores in the medial thalamus/habenula, the posterior cingulate cortex, the postcentral gyrus, and the fusiform gyrus (Table S6, Figure **1**B). The loss-related outcome error signal 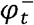 in the posterior insula was associated negatively with anhedonia scores (peak *Z*=4.73, *p*_FWEc_<.001, Table S6, Figure **1**C). No within-patients associations were observed for reward-related signals.

### 3.4 No effects of SSRI on brain activity in participants with MDD

We conducted additional analyses comparing effects of the learning parameters ***δ***, ***φ*** and Q to see if there were systematic differences of brain activity related to selective serotonin reuptake inhibitors (SSRI) medication. We compared groups of 18 medicated with SSRI to 10 other patients (unmedicated or no SSRI). However, GLM analyses of expected values and prediction error did not reveal any significant group differences. No clusters survived (*p*_FWEc_<.05) a threshold of *p*_CDT_ < .001, nor a more lenient cluster-defining threshold of *p*_CDT_ < .005. Moreover, DCM parameter values of the OFC and the dACC were assessed for SSRI effects. To this end, we extracted the individual posterior means for each patient and split them into two groups (SSRI, other or no medication). However, we did not find a significant difference between the patients taking SSRIs and others in the OFC (*t*(26)=-1.44, *p*=0.162) nor the dACC (*t*(26)=-0.08, *p*=0.936). Note that one patient was excluded form this analysis as their medication history was not disclosed.

## 4 Discussion

In this study, we used a combination of computational modeling, fMRI and connectivity analysis to study reward and loss processing in adolescent MDD. We demonstrated that adolescent MDD is associated with (a) aberrant gain control in the OFC during loss feedback processing, (b) anhedonia-related reduction of representation of loss outcomes in the posterior insula and habenula, but we found no evidence for aberrant reward prediction error processing in the ventral striatum and the medial prefrontal cortex. Thus, the present work provides novel insights into the neurobiological foundation of altered learning mechanisms in loss avoidance in early onset MDD.

Our computational modeling approach revealed differences in behavioral adaptation that were reflected in the learning rate to update one’s belief about prospective receipt of reward and loss. By testing a series of behavioral models, we showed that (a) participants with MDD adapted their instrumental vigor according to a simpler learning model with a single learning rate for reward and loss and (b) that they also updated the expected values slower than controls. While this does not indicate a different learning mechanism in MDD per se, it suggests that instrumental behavior does not rely on differential update rates to adapt behavior for approaching reward and avoiding loss. The speed of learning depends on the perceived volatility of the environment(34), which has been suggested to be affected in anhedonia and anxiety(35), the latter being a highly prevalent comorbidity in depression(36). Reduced learning and updating of one’s belief system and hence an inability to update and hold an appropriate structure of possible aversive outcomes might provide the basis for a biased evaluation of the environment.

On the neural level, we linked these differences in value representation updates derived from the computational model to neural *feedback* processing. While in controls OFC activity was related to an outcome error signal across task conditions, this was absent in participants with MDD during loss processing. Effective connectivity analysis further showed that this effect was primarily driven by aberrant gain control in the OFC, specifically the sensitivity tuning in varying magnitude contexts. The gain in the OFC is modulated by various neurotransmitters(37) but there is evidence that dopamine plays a role in learning approach and avoidance behavior(9) supported by dense dopaminergic projections to the OFC(38). This gain control might be critical to modulate the activity of the OFC during error processing in reward(39) and loss(11) contexts, updating neural representations of value(40) and maintaining a representation of the task structure(41). The latter entails updating an estimate of certainty for a specific outcome, that seems to fail in adolescent MDD when evaluating unexpected outcomes in avoidance learning. Strikingly, we found a significant negative association between the learning rate and the gain control of the OFC during loss processing. Although altered OFC activity and connectivity for aversive stimuli has been reported in various paradigms in adults with MDD(42, 43), the few studies investigating adult MDD with the MID task did not report changes in loss-related processing(44, 45). On the other hand, a longitudinal study in adolescents at risk for MDD(19) showed that loss-related OFC-insula connectivity predicted depressive symptoms nine months later. This suggests that altered OFC function during loss-processing could be specific for early-onset depression. However more research on OFC connectivity changes over the course of the disorder from adolescence into adulthood is needed. In accordance with Jin et al.(19), we also showed that decreased loss error signaling in the posterior insula was related to anhedonia. While the encoding of magnitude-modulated loss PE signals did not significantly differ between groups, a within-patients analysis revealed that BOLD responses related to loss PE in a cluster comprising the medial thalamus and habenula were significantly negatively associated with anhedonia. Previous work has implicated impaired habenula function and morphology in depression and anhedonia(6, 46). Thus, altered loss processing could reflect an important factor that contributes to increased susceptibility to adolescent MDD.

Recent computational accounts on depression suggest that it is related to an aberrant cognitive prior that underlies negative bias in evaluation of the state of the environment(47). An overgeneralization of one’s own states might eventually lead to helplessness behavior, where negative outcomes are associated with poor performance and failure of oneself, and positive outcomes are regarded as mere random events. Our results could indicate that a negative prior about the outcome is not updated due to dysfunction in the OFC, and this might contribute to maintaining a negative bias and a feeling of loss of control over outcomes. The latter is consistent with the differences of negative arousal derived from the postscan ratings, where participants with MDD expressed more relief (i.e. more deactivation) in rewarding outcomes and more fear (i.e. more activation) during loss outcomes. However, unlike in adult depression(48), computational modeling did not indicate that this higher range of negative arousal significantly affected response vigor in MDD.

Contrary to our hypothesis, we did not find any evidence of impact of depression on PE processing in rewarding contexts as previous studies in adult MDD(6, 12, 13). Here, we postulate that two factors could have led to this null finding. First, in our MID task participants did not have to learn anything to perform well. This design was employed to minimize confounds of (a) brain maturation and development within participant groups(49) and (b) diagnosis(50) on learning performance, which could be difficult to disentangle in more complex learning paradigms. Nevertheless, our results are in concordance with previous findings of intact reward PE signaling in a non-learning task in adult MDD(51). Second, there is evidence that impairments of reward PE signaling are related to the number of depressive episodes across life-time(6). This might explain the results in participants with an early onset as in our study and could indicate that previous reports of impaired reward PE signaling errors are related to the chronicity of the disorder.

It has to be considered that the majority of participants with depression were receiving antidepressive medication(Table 1). It is possible that intake of SSRIs might have affected error signaling and learning of cue-outcome associations(52). Based on this assumption, we would expect blunting of reward responses due to the administration of SSRIs. However, additional control analyses of brain activity and connectivity comparing participants with MDD with (n=18) and without (n=10) SSRI-intake revealed no significant effect. Although we cannot fully rule out that medication had an effect based on these rather small subsamples, we consider it unlikely that this was the case in this study. Nevertheless, the different medication status of patients should be considered when interepreting the results and further investigation of effects of SSRIs in depression is warranted. Moreover, we note that it is not possible to make causal inferences using a cross-sectional design, and future work should assess larger, longitudinal samples to shed light on the neural mechanisms that could give rise to MDD in adolescence.

In conclusion, this is the first study to show that adolescent MDD is associated with specific impairments of error processing in loss avoidance contexts, whereas reward sensitivity is intact. Given the critical role of evaluating an action that led to an unexpected aversive outcome, this deficit could be directly related to severe difficulties in decision-making and in social life and by contributing to the development and persistence of a negative bias in depression. Our study provides a first important step towards identifiying computational mechanisms in adolescent MDD and paves the way for establishing computational assays(53) that will facilitate the translation into clinical practice.

## Data Availability

All relevant anonymised data and code used to generate results are available from the authors on request in accordance with the requirements of the cantonal ethics board.

## 5 Acknowledgements

We thank Plamina Dimanova, Nada Frei, Dr. Noemi Baumgartner, Mona Albermann, Kristin Nalani, Paola Keller, Desiree Thommen, Luana Signer, and Dr. Philipp Stämpfli for technical and clinical assistance.

## References

1. Lopez AD, Mathers CD, Ezzati M, Jamison DT, Murray CJ (2006): Global burden of disease and risk factors. The World Bank.

2. Avenevoli S, Swendsen J, He J-P, Burstein M, Merikangas KR (2015): Major depression in the National Comorbidity Survey–Adolescent Supplement: prevalence, correlates, and treatment. Journal of the American Academy of Child & Adolescent Psychiatry. 54:37-44. e32.

3. Kieling C, Adewuya A, Fisher HL, Karmacharya R, Kohrt BA, Swartz JR, et al. (2019): Identifying depression early in adolescence. The Lancet Child & Adolescent Health. 3:211–213.

4. Cha CB, Franz PJ, M. Guzmán E, Glenn CR, Kleiman EM, Nock MK (2018): Annual Research Review: Suicide among youth–epidemiology,(potential) etiology, and treatment. Journal of Child Psychology and psychiatry. 59:460–482.

5. Chen C, Takahashi T, Nakagawa S, Inoue T, Kusumi I (2015): Reinforcement learning in depression: a review of computational research. Neuroscience & Biobehavioral Reviews. 55:247–267.

6. Kumar P, Goer F, Murray L, Dillon DG, Beltzer ML, Cohen AL, et al. (2018): Impaired reward prediction error encoding and striatal-midbrain connectivity in depression. Neuropsychopharmacology. 43:1581–1588.

7. Rescorla RA, Wagner AR (1972): A theory of Pavlovian conditioning: Variations in the effectiveness of reinforcement and nonreinforcement. Classical conditioning II: Current research and theory. 2:64–99.

8. Sutton RS (1988): Learning to predict by the methods of temporal differences. Machine learning. 3:9–44.

9. Palminteri S, Justo D, Jauffret C, Pavlicek B, Dauta A, Delmaire C, et al. (2012): Critical roles for anterior insula and dorsal striatum in punishment-based avoidance learning. Neuron. 76:998–1009.

10. Knutson B, Fong GW, Bennett SM, Adams CM, Hommer D (2003): A region of mesial prefrontal cortex tracks monetarily rewarding outcomes: characterization with rapid event-related fMRI. Neuroimage. 18:263–272.

11. Taylor SF, Martis B, Fitzgerald KD, Welsh RC, Abelson JL, Liberzon I, et al. (2006): Medial frontal cortex activity and loss-related responses to errors. Journal of Neuroscience. 26:4063–4070.

12. Gradin VB, Kumar P, Waiter G, Ahearn T, Stickle C, Milders M, et al. (2011): Expected value and prediction error abnormalities in depression and schizophrenia. Brain. 134:1751–1764.

13. Kumar P, Waiter G, Ahearn T, Milders M, Reid I, Steele J (2008): Abnormal temporal difference reward-learning signals in major depression. Brain. 131:2084–2093.

14. Nelson BD, Perlman G, Klein DN, Kotov R, Hajcak G (2016): Blunted neural response to rewards as a prospective predictor of the development of depression in adolescent girls. American Journal of Psychiatry. 173:1223–1230.

15. Stringaris A, Vidal-Ribas Belil P, Artiges E, Lemaitre H, Gollier-Briant F, Wolke S, et al. (2015): The brain’s response to reward anticipation and depression in adolescence: dimensionality, specificity, and longitudinal predictions in a community-based sample. American Journal of Psychiatry. 172:1215–1223.

16. Sharp C, Kim S, Herman L, Pane H, Reuter T, Strathearn L (2014): Major depression in mothers predicts reduced ventral striatum activation in adolescent female offspring with and without depression. Journal of abnormal psychology. 123:298.

17. Luking KR, Pagliaccio D, Luby JL, Barch DM (2016): Reward processing and risk for depression across development. Trends in cognitive sciences. 20:456–468.

18. Luking KR, Pagliaccio D, Luby JL, Barch DM (2016): Depression risk predicts blunted neural responses to gains and enhanced responses to losses in healthy children. Journal of the American Academy of Child & Adolescent Psychiatry. 55:328–337.

19. Jin J, Narayanan A, Perlman G, Luking K, DeLorenzo C, Hajcak G, et al. (2017): Orbitofrontal cortex activity and connectivity predict future depression symptoms in adolescence. Biological Psychiatry: Cognitive Neuroscience and Neuroimaging. 2:610–618.

20. Scholl J, Klein-Flügge M (2018): Understanding psychiatric disorder by capturing ecologically relevant features of learning and decision-making. Behavioural brain research. 355:56–75.

21. Knutson B, Westdorp A, Kaiser E, Hommer D (2000): FMRI visualization of brain activity during a monetary incentive delay task. Neuroimage. 12:20–27.

22. Der-Avakian A, Markou A (2012): The neurobiology of anhedonia and other reward-related deficits. Trends in neurosciences. 35:68–77.

23. Niv Y, Daw ND, Joel D, Dayan P (2007): Tonic dopamine: opportunity costs and the control of response vigor. Psychopharmacology. 191:507–520.

24. Kaufman J, Birmaher B, Brent D, Rao U, Flynn C, Moreci P, et al. (1997): Schedule for affective disorders and schizophrenia for school-age children-present and lifetime version (K-SADS-PL): initial reliability and validity data. Journal of the American Academy of Child & Adolescent Psychiatry. 36:980–988.

25. Sheehan DV, Sheehan KH, Shytle RD, Janavs J, Bannon Y, Rogers JE, et al. (2010): Reliability and validity of the mini international neuropsychiatric interview for children and adolescents (MINI-KID). The Journal of clinical psychiatry.

26. Stiensmeier-Pelster J, Braune-Krickau M, Schürmann M, Duda K Dikj - Depressionsinventar für Kinder und Jugendliche.

27. Oldham S, Murawski C, Fornito A, Youssef G, Yücel M, Lorenzetti V (2018): The anticipation and outcome phases of reward and loss processing: A neuroimaging meta□analysis of the monetary incentive delay task. Human brain mapping. 39:3398–3418.

28. Bayer HM, Glimcher PW (2005): Midbrain dopamine neurons encode a quantitative reward prediction error signal. Neuron. 47:129–141.

29. Rolls ET, Huang C-C, Lin C-P, Feng J, Joliot M (2020): Automated anatomical labelling atlas 3. NeuroImage. 206:116189.

30. Friston KJ, Harrison L, Penny W (2003): Dynamic causal modelling. Neuroimage. 19:1273–1302.

31. Zeidman P, Jafarian A, Corbin N, Seghier ML, Razi A, Price CJ, et al. (2019): A guide to group effective connectivity analysis, part 1: First level analysis with DCM for fMRI. NeuroImage.

32. Hauser TU, Iannaccone R, Walitza S, Brandeis D, Brem S (2015): Cognitive flexibility in adolescence: Neural and behavioral mechanisms of reward prediction error processing in adaptive decision making during development. Neuroimage. 104:347–354.

33. Friston KJ, Litvak V, Oswal A, Razi A, Stephan KE, Van Wijk BC, et al. (2016): Bayesian model reduction and empirical Bayes for group (DCM) studies. Neuroimage. 128:413–431.

34. Behrens TE, Woolrich MW, Walton ME, Rushworth MF (2007): Learning the value of information in an uncertain world. Nature neuroscience. 10:1214–1221.

35. Bishop SJ, Gagne C (2018): Anxiety, depression, and decision making: a computational perspective. Annual review of neuroscience.

36. Häberling I, Baumgartner N, Emery S, Keller P, Strumberger M, Nalani K, et al. (2019): Anxious depression as a clinically relevant subtype of pediatric major depressive disorder. Journal of Neural Transmission. 126:1217–1230.

37. Robbins TW, Arnsten AF (2009): The neuropsychopharmacology of fronto-executive function: monoaminergic modulation. Annual review of neuroscience. 32:267–287.

38. Kahnt T, Tobler PN (2017): Dopamine modulates the functional organization of the orbitofrontal cortex. Journal of Neuroscience. 37:1493–1504.

39. Ramnani N, Elliott R, Athwal B, Passingham R (2004): Prediction error for free monetary reward in the human prefrontal cortex. Neuroimage. 23:777–786.

40. Sul JH, Kim H, Huh N, Lee D, Jung MW (2010): Distinct roles of rodent orbitofrontal and medial prefrontal cortex in decision making. Neuron. 66:449–460.

41. Wilson RC, Takahashi YK, Schoenbaum G, Niv Y (2014): Orbitofrontal cortex as a cognitive map of task space. Neuron. 81:267–279.

42. Gao Q, Zou K, He Z, Sun X, Chen H (2016): Causal connectivity alterations of cortical-subcortical circuit anchored on reduced hemodynamic response brain regions in first-episode drug-naïve major depressive disorder. Scientific reports. 6:1–12.

43. Schiller CE, Minkel J, Smoski MJ, Dichter GS (2013): Remitted major depression is characterized by reduced prefrontal cortex reactivity to reward loss. Journal of affective disorders. 151:756–762.

44. Ubl B, Kuehner C, Kirsch P, Ruttorf M, Diener C, Flor H (2015): Altered neural reward and loss processing and prediction error signalling in depression. Social cognitive and affective neuroscience. 10:1102–1112.

45. Pizzagalli DA, Holmes AJ, Dillon DG, Goetz EL, Birk JL, Bogdan R, et al. (2009): Reduced caudate and nucleus accumbens response to rewards in unmedicated individuals with major depressive disorder. American Journal of Psychiatry. 166:702–710.

46. Lawson R, Nord C, Seymour B, Thomas D, Dayan P, Pilling S, et al. (2017): Disrupted habenula function in major depression. Molecular psychiatry. 22:202–208.

47. Clark JE, Watson S, Friston KJ (2018): What is mood? A computational perspective. Psychological Medicine. 48:2277–2284.

48. Steele J, Kumar P, Ebmeier KP (2007): Blunted response to feedback information in depressive illness. Brain. 130:2367–2374.

49. Nussenbaum K, Hartley CA (2019): Reinforcement learning across development: What insights can we draw from a decade of research? Developmental cognitive neuroscience.100733.

50. Snyder HR (2013): Major depressive disorder is associated with broad impairments on neuropsychological measures of executive function: a meta-analysis and review. Psychological bulletin. 139:81.

51. Rutledge RB, Moutoussis M, Smittenaar P, Zeidman P, Taylor T, Hrynkiewicz L, et al. (2017): Association of neural and emotional impacts of reward prediction errors with major depression. JAMA psychiatry. 74:790–797.

52. McCabe C, Mishor Z, Cowen PJ, Harmer CJ (2010): Diminished neural processing of aversive and rewarding stimuli during selective serotonin reuptake inhibitor treatment. Biological psychiatry. 67:439–445.

53. Stephan KE, Mathys C (2014): Computational approaches to psychiatry. Current opinion in neurobiology. 25:85–92.

