## Supplement for "Maladaptive avoidance learning in the orbitofrontal cortex in adolescents with major depression"

### Supplementary Methods

#### Behavioral raw data analysis

Response data were log-transformed to achieve a more normally distributed data set for data analysis. Data that deviated more than three standard deviations from the respective mean per condition and per subject were excluded from the analysis (2%). For a conventional analysis, we averaged the logRTs for each subject and each condition and performed an ANOVA to assess effects of condition, group, and their interaction.

The post-scan ratings were mean-centered within subject and rotated to obtain measures of positive and negative arousal for each trial type (high/low reward, high/low loss, neutral) as they increase interpretability in terms of approach and avoidance behavior (1). To this end, we calculated negative and positive arousal for each outcome according to the equations:

$$\begin{aligned} Positive Arousal=\frac{\left( Arousal +\mathrm{Valence} \right)}{\sqrt{2}}\#\left( 1 \right) \end{aligned}$$

$$\begin{aligned} Negative Arousal=\frac{\left( Arousal-V\mathrm{alence} \right)}{\sqrt{2}}\#\left( 2 \right) \end{aligned}$$

We had to exclude four controls from this behavioral analysis as they did not complete the ratings due to time constraints. We performed an ANOVA with condition and group as fixed factor. The significance level for all statistical tests of the behavioral analyses was p < 0.05, two-tailed.

#### Behavioral modeling

##### Computational learning algorithm

To assess error processing in our participants, we fitted a Rescorla-Wagner learning model (2) to the response time data of the participants. Influential work established that activity in brain regions modulated by dopamine correlate with an *expected value* $Q_{t}$ and *reward prediction errors* $\delta_{t}$ (3, 4). Later, studies found that these signals play critical roles in approach and avoidance learning in humans (5, 6). Herein, we defined two different signals, based on the current cue in our task (gain reward, avoid loss) to model and disentangle effects of loss and reward. At cue presentation, the model assumes that each participant builds up an expected value based on their subjective probability to win or lose:

$$\begin{aligned} Q_{t}^{+}=C_{t}{\cdot v}_{t}\#\left( 3 \right) \end{aligned}$$

$$\begin{aligned} Q_{t}^{-}=C_{t}{\cdot(1-v}_{t})\#\left( 4 \right) \end{aligned}$$

Here, $Q_{t}^{+}$ represents an expected reward, whereas $Q_{t}^{-}$ represents an expected loss, dependent on the subjective probability for a reward $v_{t}$ and loss $(1- v_{t})$ and the possible outcome $C_{t}$. It is crucial to reinforcement learning models that the expected values are updated on the difference between the expected value and the outcome in each observation which is the prediction error. During receipt or omission of reward or loss respectively, prediction errors are thought to be teaching signals that enable the adaptation of future behavior to optimize outcome and continue to be computed even when behavior is already highly trained (7). Importantly, such an update mechanism allows for establishing and tracking the value in environments, where obtaining a reward is probabilistic. The probability to encounter a miss was $P\left( Hit \right)=1-P\left( Hit \right)\approx33\%$ and ensured across participants by adjusting the difficulty based on the performance history using an adaptive algorithm. Conventional reward ($\delta_{t}^{+}$) and loss ($\delta_{t}^{-}$) prediction error signals were calculated as the difference between the expected value and the trial outcome (0, ±1, ±4 CHF):

$$\begin{aligned} \delta_{t}^{+}=\left\{ \begin{matrix} R_{t}-Q_{t}^{+} & C_{t}>0 \\ 0 & C_{t}\leq0 \end{matrix} \right.\#\left( 5 \right) \end{aligned}$$

$$\begin{aligned} \delta_{t}^{-}=\left\{ \begin{matrix} Q_{t}^{-}-R_{t} & C_{t}<0 \\ 0 & C_{t}\geq0 \end{matrix} \right.\#\left( 6 \right) \end{aligned}$$

As in previous work (8), the signal for updating future predictions $\varphi$ was scaled by the magnitude of the experienced outcome, such that the update was independent of the experienced reward/loss value:

$$\begin{aligned} \varphi_{t}^{+}=\left\{ \begin{matrix} \frac{R_{t}}{C_{t}}-v_{t} & C_{t}\neq0 \\ 0 & C_{t}=0 \end{matrix} \right.\#\left( 7 \right) \end{aligned}$$

$$\begin{aligned} \varphi_{t}^{-}=\left\{ \begin{matrix} {(1-v}_{t})-\frac{R_{t}}{C_{t}} & C_{t}\neq0 \\ 0 & C_{t}=0 \end{matrix} \right.\#\left( 8 \right) \end{aligned}$$

By this definition, *outcome errors* $\varphi_{t}^{+}$ and $\varphi_{t}^{-}$ reflect “surprise” signals that can be used for learning stimulus-outcome associations in either reward or loss context, whose size depends solely on the reinforcement history and not value. In other words, this update mechanism assumes that the participants change their belief about future hits and misses equally across cue magnitudes.

The prediction error is weighted by a *learning rate*$\alpha$, which determines the step size of the belief adjustment to hit the target in a future trial. As behavioral adaptation might occur with different rates in controls and patients in reward and loss contexts, respectively, we defined two different reinforcement learning models: one model with a single learning rate for both updates (rw1, eq. 9), one with separate learning rates for reward and loss updates (rw2, eq. 10). The update rules for the hit probability in the subsequent trial in the respective models were therefore:

Rescorla-Wagner Model 1 (rw1)

$$\begin{aligned} v_{t+1}=\left\{ \begin{matrix} v_{t}+\alpha\cdot\varphi_{t} & C_{t}\neq0 \\ v_{t} & C_{t}=0 \end{matrix} \right.\#(9) \end{aligned}$$

Rescorla-Wagner Model 2 (rw2)

$$\begin{aligned} v_{t+1}=\left\{ \begin{matrix} v_{t}+\alpha^{+}\cdot\varphi_{t}^{+} & C_{t}>0 \\ v_{t} & C_{t}=0 \\ v_{t}+\alpha^{-}\cdot\varphi_{t}^{-} & C_{t}<0 \end{matrix} \right. \#(10) \end{aligned}$$

where $\varphi_{t}$ denotes the overall signed prediction error in both contexts.

In addition, *average reward and loss* at each trial was defined as:

$$\begin{aligned} \bar{R}_{t}=\left\{ \begin{matrix} \bar{R}_{t-1}+\alpha\cdot(R_{t-1}-\bar{R}_{t-1}) & C_{t}>0 \\ \bar{R}_{t-1} & C_{t}\leq0 \end{matrix} \right.\#\left( 11 \right) \end{aligned}$$

$$\begin{aligned} \bar{L}_{t}=\left\{ \begin{matrix} \bar{L}_{t-1}+\alpha\cdot({|L}_{t-1}|-\bar{L}_{t-1}) & C_{t}<0 \\ \bar{L}_{t-1} & C_{t}\geq0 \end{matrix} \right.\#\left( 12 \right) \end{aligned}$$

where $R_{t}$ and $L_{t}$ represent the actual and $\bar{R}_{t}$ and $\bar{L}_{t}$ the average reward or loss at trial t (9, see below). For the rw2 model, separate learning rates $\alpha$ were used for rewards and losses.

##### Response model

Expected values (10) and prediction errors (11) have been shown to be able to modulate motivated behavior in terms of the response vigor. Thus, to investigate performance adaptation for varying expected values and prediction errors across the fMRI task, we used the trajectories resulting from the learning model to generate trial-by-trial predictions of logRTs which served as a proxy for their motivation. We compared five different plausible response models: All models assume that the logRT is a linear combination of individual task-related parameters and a constant term. Each response model included (a) the expected values, as they are thought to increase response vigor (10), and (b) a linear function *g,* that captured any systematic increase/decrease in reaction times throughout the task. In total, five models with varying factors affecting response vigor were created and compared using a Bayesian model selection procedure. Response model M1 (equation 13) emphasized average reward and loss rates as proposed by previous work (9, 12). Reward and loss prediction errors are associated with dopaminergic activity and improved performance (11). As loss prediction errors are likely signaled via a different mechanism (13), in another response model reward and loss were allowed to influence the reponse vigor separately (M2, equation 14). Lastly, the unsigned expected values or prediction errors, reflecting cue salience and novelty, respectively, have been shown to modulate dopaminergic activity (14). Thus, we tested models in which combinations of (M3, equation 15) cue salience and novelty, (M4, equation 16) signed expected values, or (M5, equation 17) signed prediction errors could influence response vigor. $PostError$ represents a vector of trials after a miss, $Rep$ represents a vector of successive presentation of equal cues, and $\zeta$ represents Gaussian noise.

*Response model M1:*

$$\begin{aligned} {\log\left( RT \right)}^{t}=\beta_{0}+\beta_{1}\cdot{|Q}_{t}^{-}|+\beta_{2}\cdot Q_{t}^{+}+\beta_{3}\cdot\bar{R}_{t-1}+\beta_{4}\cdot\bar{L}_{t-1}+ \\ + \beta_{5}\cdot{PostError}_{t}+\beta_{6}\cdot{Rep}_{t}+\beta_{7}\cdot g\left( t \right)+\zeta_{t}\#\left( 13 \right) \end{aligned}$$

*Response model M2:*

$$\begin{aligned} {\log\left( RT \right)}^{t}=\beta_{0}+\beta_{1}\cdot{|Q}_{t}^{-}|+\beta_{2}\cdot Q_{t}^{+}+{\beta_{3}\cdot\delta_{t-1}^{-}+\beta}_{4}\cdot\delta_{t-1}^{+}+\beta_{5}\cdot g\left( t \right)+ \zeta_{t}\#\left( 14 \right) \end{aligned}$$

*Response model M3:*

$$\begin{aligned} {\log\left( RT \right)}^{t}=\beta_{0}+\beta_{1}\cdot{|Q}_{t}|+\beta_{2}\cdot{|\delta}_{t-1}|+\beta_{3}\cdot{PostError}_{t}+\beta_{4}\cdot g\left( t \right)+ \zeta_{t}\#\left( 15 \right) \end{aligned}$$

*Response model M4:*

$$\begin{aligned} {\log\left( RT \right)}^{t}=\beta_{0}+\beta_{1}\cdot{|Q}_{t}^{-}|+\beta_{2}\cdot Q_{t}^{+}+\beta_{3}\cdot{|\delta}_{t-1}|+\beta_{4}\cdot{PostError}_{t}+\beta_{5}\cdot g\left( t \right)+ \zeta_{t}\#\left( 16 \right) \end{aligned}$$

*Response model M5:*

$$\begin{aligned} {\log\left( RT \right)}^{t}=\beta_{0}+\beta_{1}\cdot{|Q}_{t}|+{\beta_{2}\cdot\delta_{t-1}^{-}+\beta}_{3}\cdot\delta_{t-1}^{+}+\beta_{4}\cdot g\left( t \right)+ \zeta_{t}\#\left( 17 \right) \end{aligned}$$

##### Model fitting

All behavioral models were fitted using the TNU Algorithms for Psychiatry-Advancing Science (TAPAS, <http://www.translationalneuromodeling.org/tapas>) HGF Toolbox 5.3. Trials without response were omitted during model fitting; priors are reported in Table S3. We used random-effects Bayesian Model Selection (15) to choose the best-fitting model by comparing the negative free energies. Besides the posterior probabilities, the exceedance probability (XP) of each model is reported. In a last step, we assessed group differences of reponse parameters. Note, that for the learning rate parameter we detected an outlier after visual inspection and a significant Grubb’s test. Thus, for this parameter, we performed a group comparison with and without this subject. Intercorrelations between parameters were assessed after excluding this subject for this analysis. However, as this participant was a patient and the higher learning rate might reflect a pathological process, we included this subject in further analyses. For group comparisons, we used either two-sample *t*-tests or Mann-Whitney *U*-Tests, in case the Shapiro-Wilk test indicated violation of normality assumptions in at least one of the groups. Finally, posterior predictive checks were conducted to assess the reliability of the behavioral model. For this, we averaged the logRTs of 1000 simulations with the individual parameters of the best-fitting model for each subject in TAPAS and mirrored the raw data analysis with the synthetic data.

#### Image acquisition and preprocessing

MRI recordings were conducted on an Achieva 3T scanner (Philips Medical Systems, Best, the Netherlands) using the manufacturer’s 32-channel head coil array. Functional T2*-weighted image acquisition was performed using a multi-slice echo-planar images (EPI) sequence [335volumes per session, $TR=1600ms$, $TE=35ms$,$50$slices, voxel size $=2.4\times2.4\times2.2{mm}^{3}$, matrix size $=76\times78px$, flip angle$=75^{\circ}$, gap $=0.35mm$, SENSE-factor$=2$, MB-factor $=2$]. To improve signal quality in the orbiofrontal cortex, which was of particular interest for this study, we tilted the field of view $15^{\circ}$downwards of AC-PC. The first five dummy scans were discarded. For the normalization procedure, a T1-weighted structural scan was acquired for each subject [MP-RAGE, aligned at AC-PC, flip angle$=9^{\circ}$, voxel size $=1.05\times1.05\times1.2{mm}^{3}$, field of view $=270\times253{mm}^{2}$, $170$ sagittal slices]. The functional data was first slice-time corrected, then realigned and unwarped using the B0-field map and coregistered to the T1-weighted image. The deformation fields derived from the segmentation of the T1 image were used for normalization to the Montreal Neurological Institute (MNI)-152 template space. The normalized volumes were spatially smoothed using a $6mm$ full-width-half-maximum kernel. All steps were conducted in SPM12 (7487). To account for motion artefacts during the scan, we calculated the framewise displacement (FD) across volumes (16). No subject exceeded a mean FD of 0.5mm ($M=0.18, SD= 0.08mm$), however, single volumes that exceeded a FD greater than $1mm$were censored in the ensuing analyses by including an additional binary regressor (% volumes censored per subject $M=0.92, SD= 1.90\%$).

#### fMRI GLM analysis

On the first level, the anticipation phase was modelled as a boxcar function from cue onset until button press and the feedback phase was modelled as event at outcome presentation. Expected values were used as parametric modulators for the anticipation phase and reward-/loss prediction errors for the feedback phase, respectively, for both reward and loss separately. The effect of the outcome error, i.e. the magnitude-independent error signal, was assessed in a separate GLM using one parametric modulator for outcome magnitude and one for the magnitude-independent error signals for reward and loss trials separately. The modulator for the outcome error was orthogonalized with respect to the outcome magnitude regressor, such that any shared variance between the correlating regressors was assigned to the latter. This outcome error regressor captures the deviation from the expected outcome (hit or miss) independent of the magnitude context. With this approach, we not only investigated brain regions encoding the effect of conventional reward/loss prediction errors with the multiplicative term of incentive probability $\times$ outcome, but also reveal brain regions, that code deviations from expected reward/loss outcomes across trials irrespective of magnitude. All first-level models included six realignment parameters and a binary vector for scans with $>1mm$ FD as nuisance regressor to the model. Neutral trials were modeled in a separate regressor. Finally, we applied a 1/128Hz cut-off high-pass filter to eliminate low frequency drifts.

#### DCM analysis

The goal of the DCM analysis was to identify the network dynamics related to expected value and prediction error processing in loss that give rise to the observed finding of decreased loss-related OFC activity in adolescents with MDD. In particular, we investigated whether the group effect revealed by the GLM analyses was located in the OFC or was a downstream effect of PE signaling in the insula and the anterior cingulate cortex, and how specific the deviations were to the magnitude-related PE. Thus, we set up a separate GLM model for the DCM analysis: the driving input were the outcome events which entered the inferior occipital gyrus. The regressors for the modulation on the self-connections (i.e. the context-dependent input sensitivity) comprised the magnitude-independent PE, the magnitude (high, low) and their interaction (magnitude-dependent PE, ± low PE, ± high PE), for reward and loss separately. This allowed us to disentangle different aspects attributed to the outcome and to test whether decreased orbitofrontal activity during loss outcomes is the result of error or loss-magnitude processing.

### Supplementary Results

#### Behavioral raw data analysis

A group-by-condition ANOVA for mean logRTs did not reveal a significant effect of condition, *F*(4, 305) = 0.95, *p* = 0.434, nor the interaction term, *F*(4, 305) = 0.23, *p* = 0.921, but a trend in the factor group, *F*(1, 305) = 3.532, *p* = 0.061. The number of response omissions did not differ between groups (HC: M = 5.4. SD = 2.9; MDD: M = 6.4, SD = 5.1; U = 460, *p* = 0.633), yielding comparable hit rates across conditions (Table S1). This suggests that the MID task was well balanced for both groups, showing no behavioral group differences in terms of reaction times and response omissions.

#### Model analysis of dual learning rate model

We conducted an additional analysis to compare patients’ and controls’ learning rates for the more complex dual learning rate model (rw2). However, there was no significant difference between the learning rates when using the (worse-fitting) dual learning rate model ($\alpha^{+}$ :MDD,0.047 [0.009]; controls, 0.049 [0.008]; W=391; *p*=.155; $\alpha^{-}$: MDD,0.052 [0.011]; controls, 0.054 [0.013]; W=447; *p*=.516). Moreover, there was no significant difference between the learning rates for losses and rewards in healthy controls. ($\alpha^{+}$-$\alpha^{-}$: V=187, *p*=.097).

#### Control analysis and model simulation

Posterior means of the behavioral model showed only a moderate correlation among each other (all |*r*| < 0.53 (Figure S3). We simulated behavioral data from the model parameters obtained for each participant and repeated the raw data analysis of synthetic logRTs. Note that in the raw data analysis we found a trend of faster response in the patients group. This trend was replicated in the analysis of variance with the simulated data: main effect of group, *F*(1,305) = 3.145, *p* = .078, main effect of condition, *F*(4,305) = 0.980, *p* = .418, group-by-condition interaction, *F*(4,305) = 0.251, *p* = .909. In addition, observed and simulated mean logRTs showed a strong correlation, Spearman’s $\boldsymbol{\rho}$ **=** 0.828**,** *p* < 10^-15^ . Hence, this suggests that the simulated data was comparable to the empirical data in both groups.

#### Increased negative arousal of HC and MDD in post-scan ratings

The analysis of ratings of negative arousal revealed a significant main effect of condition *F*(4, 295) = 156.04, *p* < 10^-15^, and a group-by-condition interaction, *F*(4, 295) = 7.73, *p* < 10^-5^, specifically patients showed higher negative arousal in loss (-4CHF: *p* = .018; -1 CHF: *p* <.001) and lower negative arousal in reward (+4CHF: *p* = .031; +1CHF: *p* = .011) than controls (Figure S2). Positive arousal ratings were comparable between groups and across conditions, group-by-condition: *F*(4,295) = 0.56, *p* = .693.

#### Neural correlates of outcome error ($\boldsymbol{\varphi}$) processing: main effects

We located increasing activity encoding of reward outcome errors ($\boldsymbol{\varphi}_{\boldsymbol{t}}^{\boldsymbol{+}}$) in the putamen, caudate, orbitofrontal cortex (OFC), lateral prefrontal cortex (PFC), temporal lobe, whereas activity in the insula, dorsal anterior cingulate gyrus (ACC), and ventrolateral PFC decreased (Table S5).

When processing loss (i.e. negative $\boldsymbol{\varphi}_{\boldsymbol{t}}^{\boldsymbol{-}}$), $\boldsymbol{\varphi}_{\boldsymbol{t}}^{\boldsymbol{-}}$ was negatively associated with activity in the anterior insula, dorsal ACC, ventrolateral PFC and the supramarginal gyrus. Avoiding loss (i.e. positive $\boldsymbol{\varphi}_{\boldsymbol{t}}^{\boldsymbol{-}}$) was associated with clusters in the caudate and putamen, dorsolateral PFC, superior temporal gyrus, paracentral lobule, lingual gyrus and occipital lobe (Table S5, see Figure 1 in main text).

#### Neural correlates of prediction error ($\boldsymbol{\delta}$) processing: main effects

The difference between reward magnitude and expected value ($\boldsymbol{\delta}_{\boldsymbol{t}}^{\boldsymbol{+}}$) during the feedback phase was positively associated with BOLD changes in the ventral and dorsal striatum, the ventromedial PFC, OFC, postcentral gyrus, temporal lobes, and the occipital lobe for increasing $\boldsymbol{\delta}_{\boldsymbol{t}}^{\boldsymbol{+}}$. A network containing the anterior insula, dorsomedial PFC, and ventrolateral PFC was negatively associated with $\boldsymbol{\delta}_{\boldsymbol{t}}^{\boldsymbol{+}}$ (Table S7). During loss processing, $\boldsymbol{\delta}_{\boldsymbol{t}}^{\boldsymbol{-}}$ was positively associated with activation in the caudate, putamen, middle temporal, superior frontal, middle frontal, and postcentral cortex and superior parietal lobe. In addition, decreasing $\boldsymbol{\delta}_{\boldsymbol{t}}^{\boldsymbol{-}}$ was associated with higher activation within the dorsal ACC, dorsomedial PFC, anterior insula, middle temporal gyrus, ventrolateral PFC, supramarginal gyrus, and midbrain (Table S7). We did not find any differences between patients and controls.

#### Neural correlates of expected value (Q) processing: main effects

We analysed the main effect of expected value during the anticipation phase of the task, and found a network that was positively associated with increasing Q+ comprising the ventromedial PFC, lateral PFC, ventral striatum, anterior insula, midbrain (Table S9, Figure S6). No cluster was found that was negatively associated with Q+. Furthermore, we did not find any cluster that was associated during processing the expected loss value Q- across participants. No differences between patients and controls were observed.

### Supplementary Figures

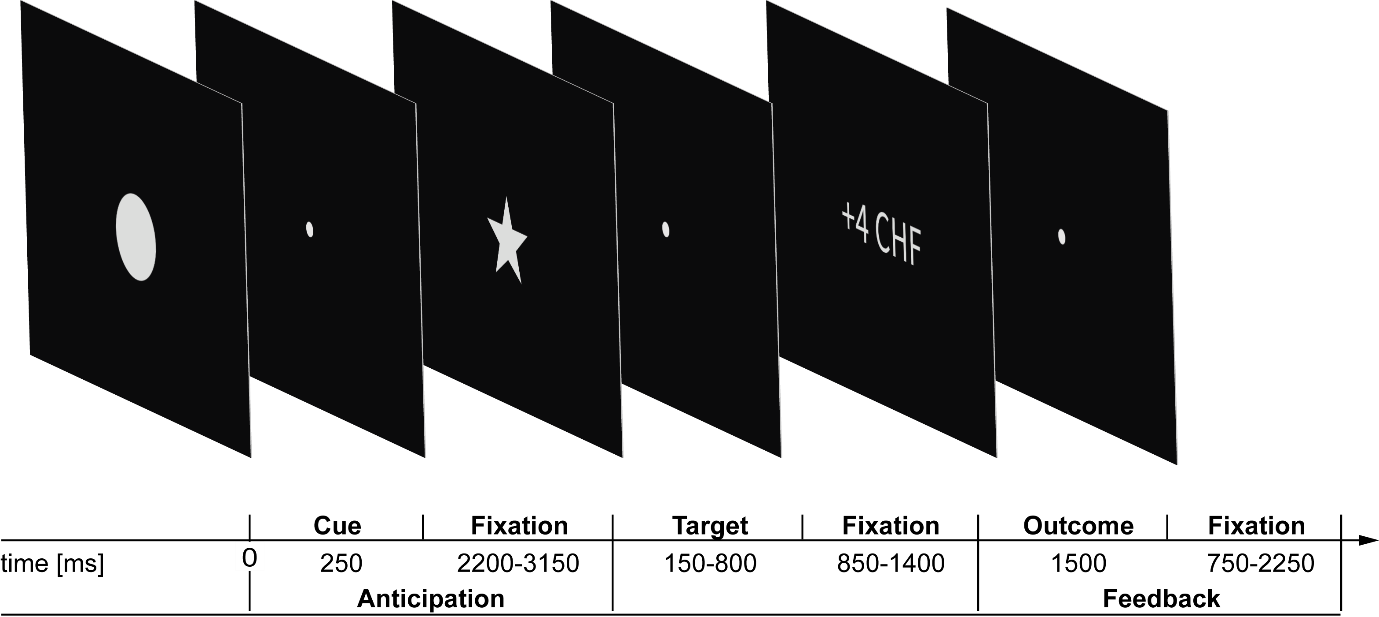

Figure S1. The monetary incentive delay (MID) task examines incentive anticipation and feedback processing. Here, the task comprised different conditions with outcomes varying in level of magnitude (low, high) and valence (reward, loss, null) that were indicated by a cue at the beginning of each trial. Participants had to respond as fast as possible to a go-signal (star symbol), which was presented after a variable delay. After a brief fixation period, participants were presented with the actual outcome of the trial. The task comprised 24 trials per cue, i.e. 120 trials in total, with an experimentally set hit rate of ≈ 66%.

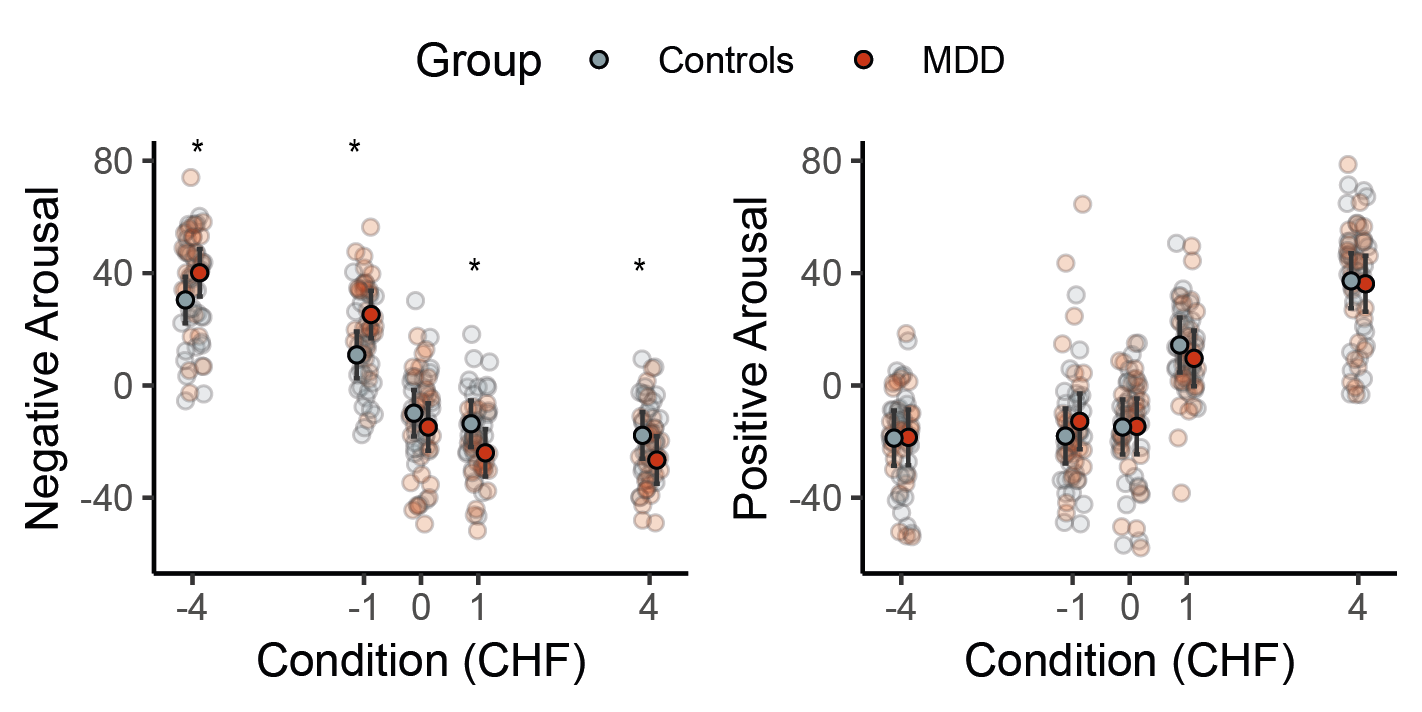

Figure S2. Subjective negative and positive arousal of outcomes. Post-scan ratings of subjective liking and arousal for each trial type was centered and rotated to obtain estimates of negative and positive arousal. Patients differed significantly on the negative arousal scale, by rating loss higher and reward lower than controls. No group difference was observed in the positive arousal scale.

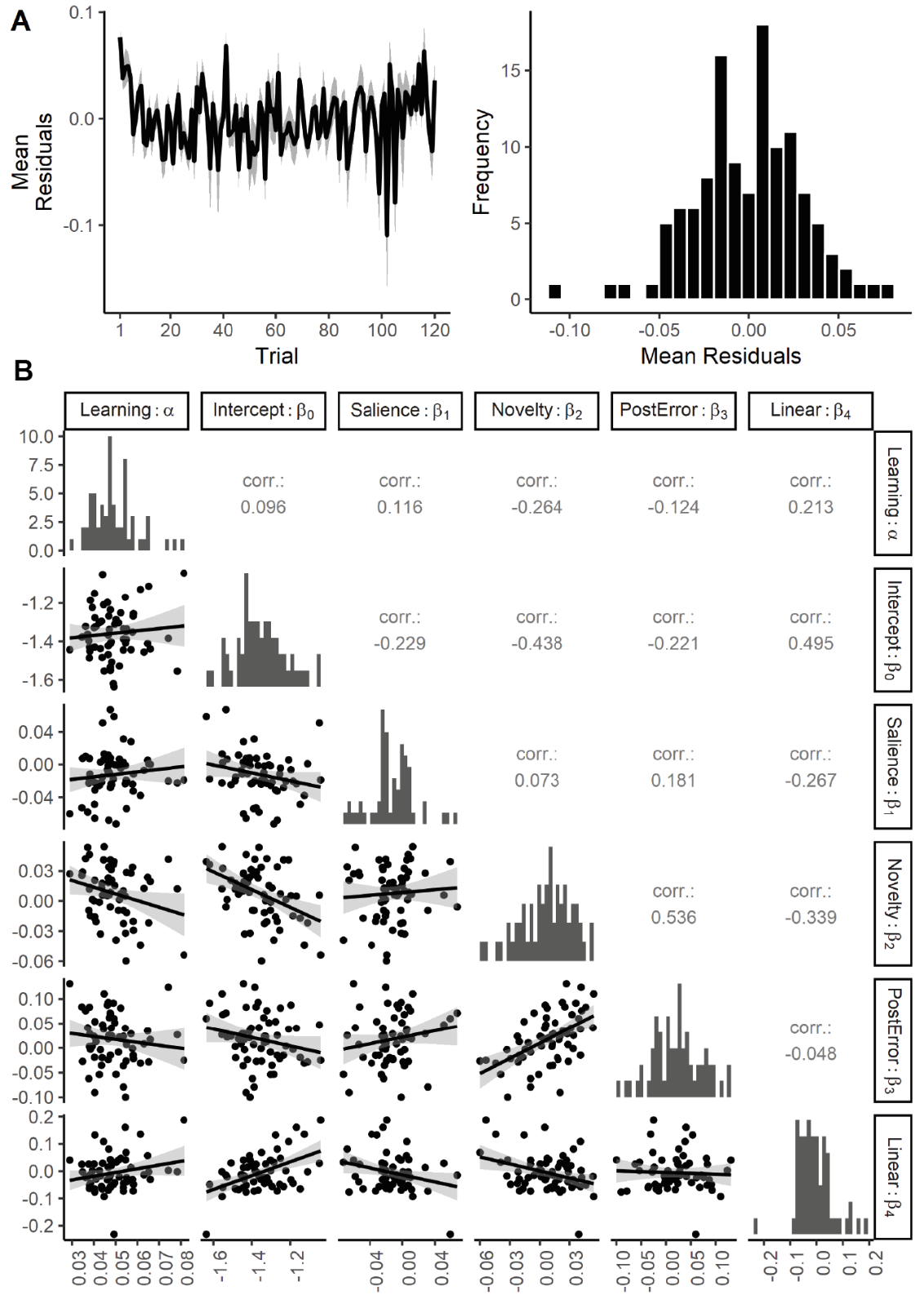

Figure S3. Assessment of the best-fitting behavioral model. (A) The distribution of the model residuals suggested that the model was able to capture the response patterns in the empirical data across participants. Shaded area indicates the SEM. (B) Intercorrelations between the parameters of the winning behavioral model. We found only small to moderate correlations between the parameters of the behavioral model. This suggests that the effects predicting the response vigor could be disentangled well in the model. Corr values are Pearson correlation coefficients. Data points represent the beta values. n=62.

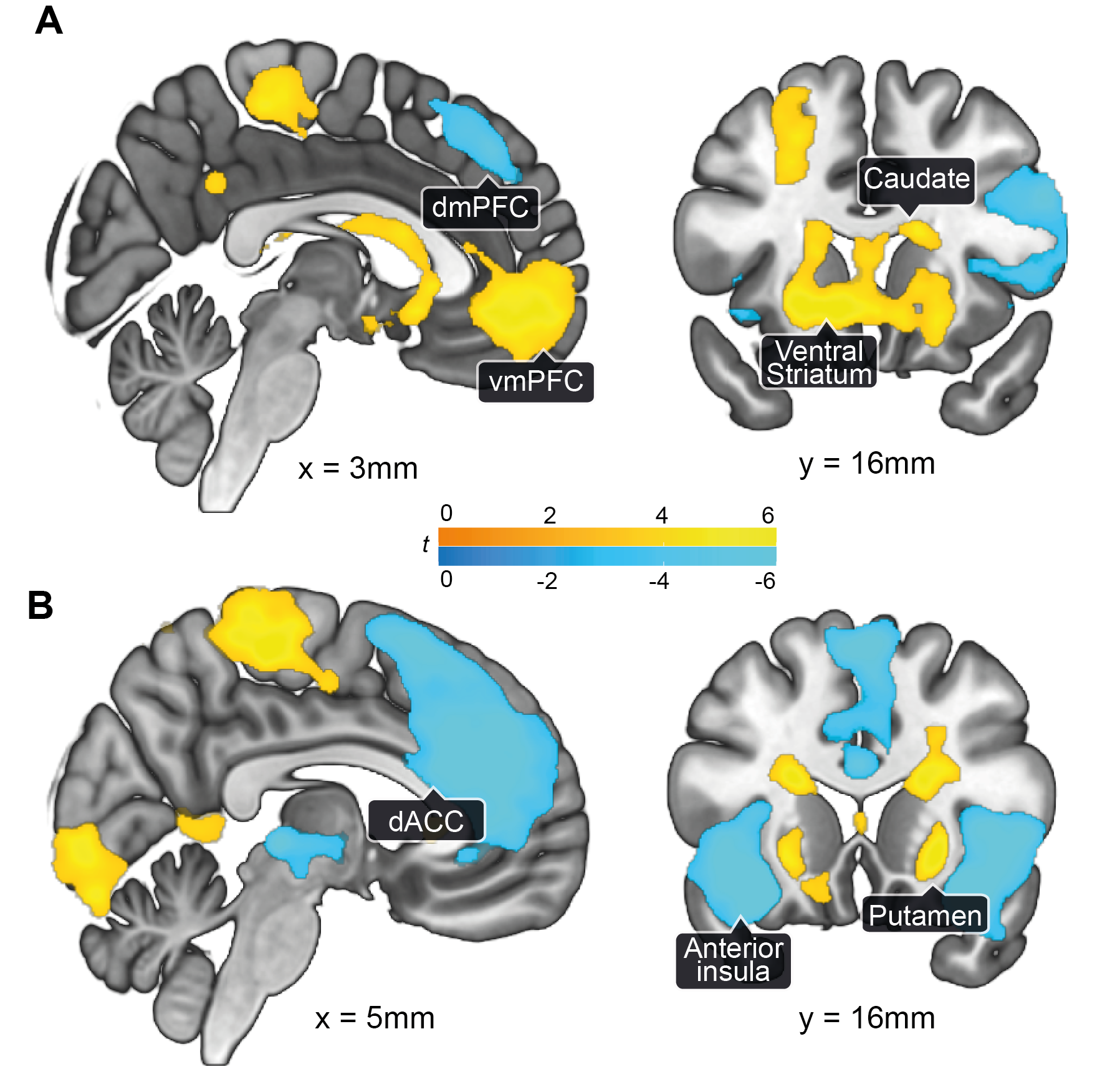

Figure S4. Activity associated with magnitude-related PE across groups. (A) The network encoding increasing (yellow) magnitude-related PE $\boldsymbol{\delta}_{\boldsymbol{t}}^{*}$ was related to activity in the ventral striatum, caudate, and the ventromedial prefrontal cortext (vmPFC). Decreasing (blue) $\boldsymbol{\delta}_{\boldsymbol{t}}^{*}$ modulated activity in the lateral prefrontal cortex and the dorsomedial PFC (dmPFPC). (B) Activity in the putamen and caudate was related to increasing (yellow) $\boldsymbol{\delta}_{\boldsymbol{t}}^{-}$, whereas activity in the insula and the medial prefrontal cortex, especially the dorsal ACC, was associated with decreasing (blue) $\boldsymbol{\delta}_{\boldsymbol{t}}^{-}$. p_FWEc_ < .05, p_CDT_ < .001, n = 63.

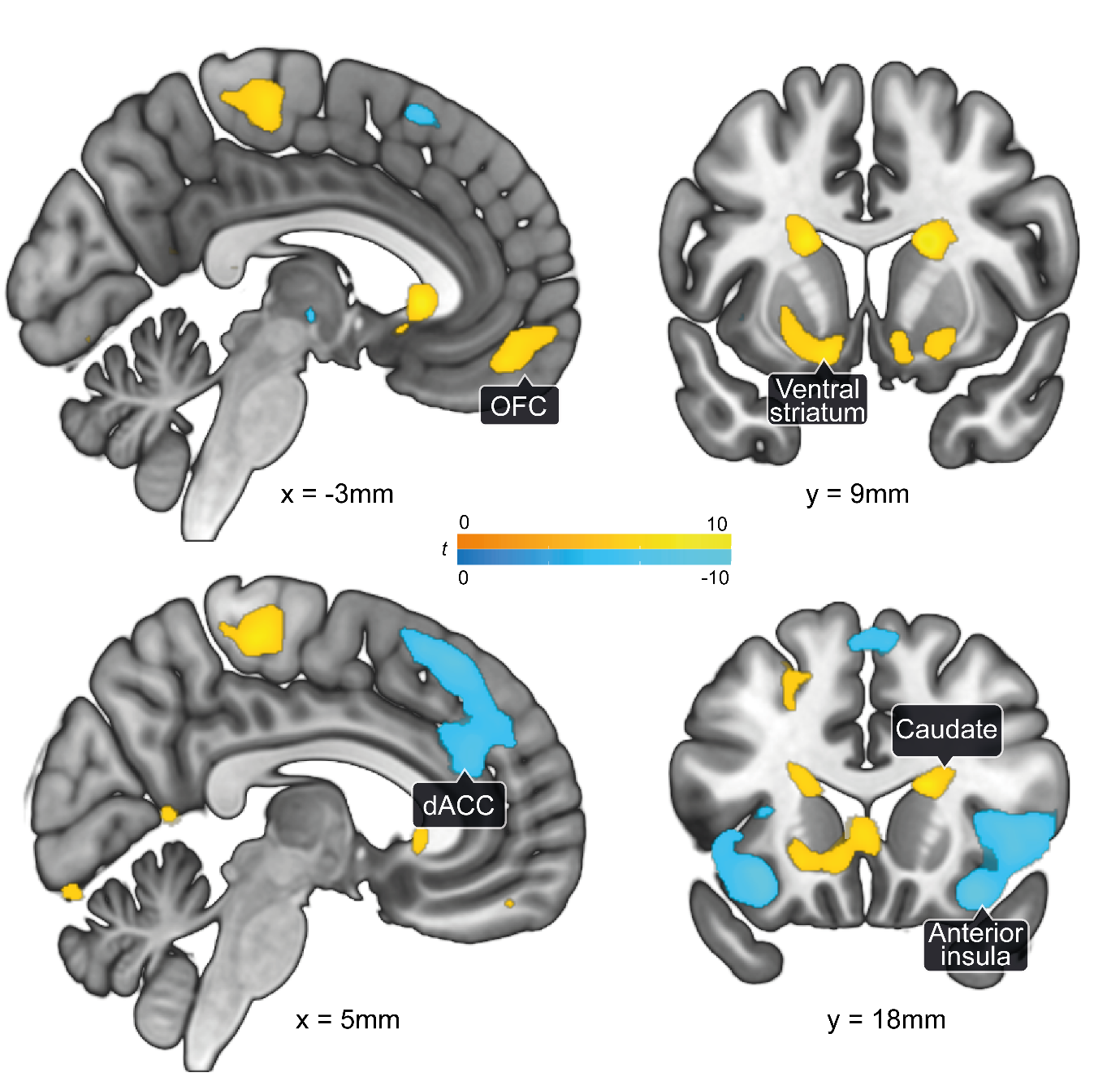

Figure S5. Activity associated with hits and misses across groups. Distinct brain activation in networks of hit (reward and loss avoidance, yellow) and miss (reward omission and loss, blue). p_FWE_ < .05, n = 63.

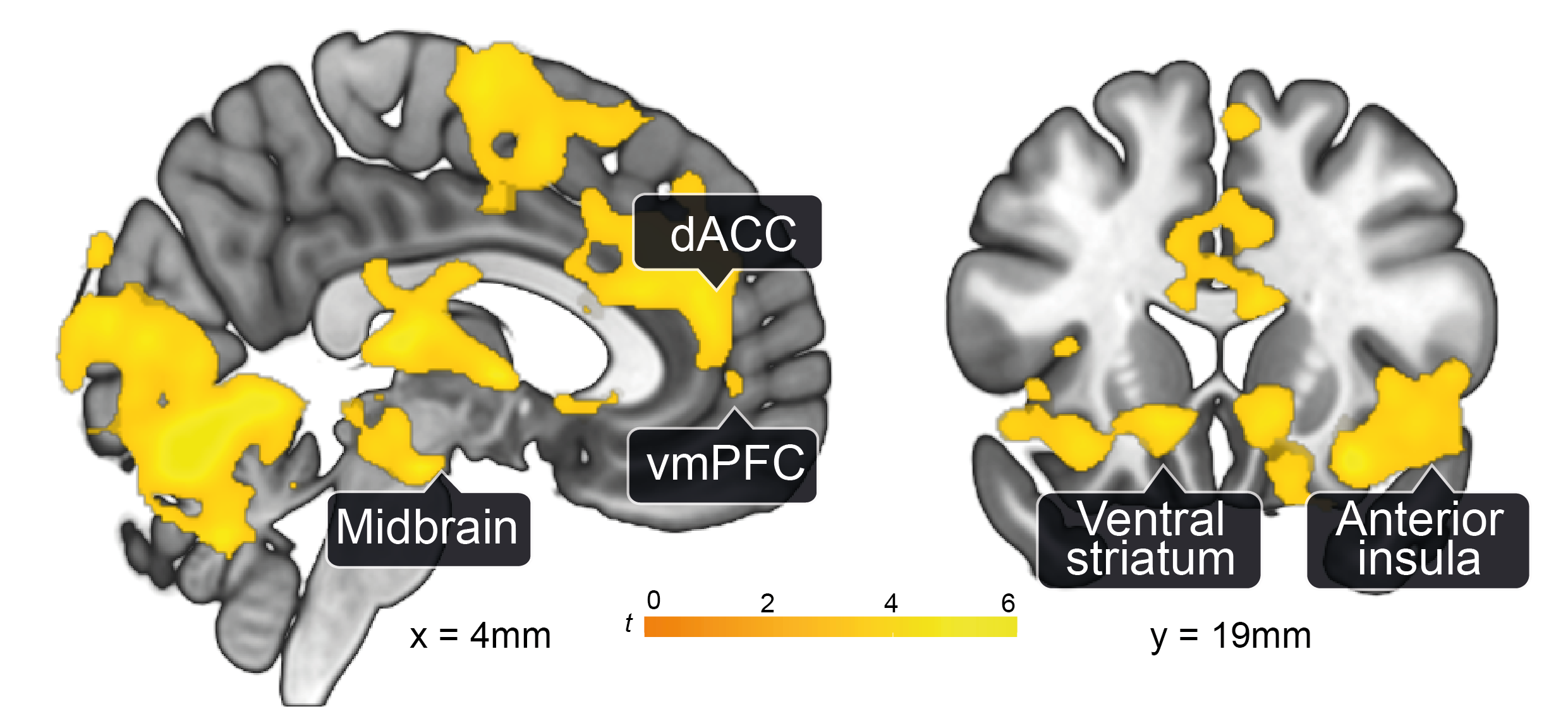

Figure S6. Activity associated with expected reward values Q+ across groups. Across both groups, an increase in expected reward values Q+ was associated with increased activation in the midbrain, the ventral striatum, the anterior insula, the dorsal anterior cingulate cortex (dACC), and the ventromedial prefrontal cortex (vmPFC). p_FWEc_ < .05, p_CDT_ < .001, n = 63.

### Supplementary Tables

| Table S1. Behavioral data of the monetary incentive delay task. Experimental manipulation ensured that a hit rate of around 66% was achieved. | | | | | | | | | | |
| --- | --- | --- | --- | --- | --- | --- | --- | --- | --- | --- |
|  | **High loss** | | **Low loss** | | **Neutral** | | **Low Gain** | | **High Gain** | |
|  | HC | MDD | HC | MDD | HC | MDD | HC | MDD | HC | MDD |
| Hit rate | 61 (9)% | 64 (10)% | 58 (12) % | 57 (12)% | - | - | 60 (11)% | 63 (13)% | 64 (10) % | 62 (12)% |
| Response time | 268 (11) ms | 259 (10) ms | 276 (16) ms | 270 (11) ms | 269 (10) ms | 265 (10) | 270 (10) ms | 261 (12) | 265 (9) ms | 254 (8)ms |
| Arousal rating | 59.4 (30.8) | 65.3 (30.7) | 46.1 (28.5) | 58.8 (25.2) | 33.7 (24.1) | 29.2 (21.5) | 51.6 (22.4) | 39.8 (26.6) | 64.9 (27.6) | 56.8 (33.8) |
| Valence rating | 13.1 (20.3) | 5.7 (9.7) | 27.4 (23.3) | 20.3 (23.4) | 44.4 (19.4) | 47.4 (21.9) | 67.7 (17.6) | 71.0 (19.3) | 86.8 (21.1) | 91.6 (12.3) |
| Negative arousal | 30.4 (18.7) | 40.1 (17.5) | 10.9 (15.7) | 25.2 (14.9) | -9.9 (15.2) | -14.8 (18.9) | -13.7 (14.3) | -24.0 (11.6) | -17.8 (15.4) | -26.6 (15.1) |
| Positive arousal | -18.8 (16.9) | -18.5 (16.9) | -18.0 (18.1) | -12.8 (23.9) | -14.8 (18.1) | -14.6 (18.3) | 14.4 (12.9) | 9.6 (18.0) | 37.2 (21.6) | 36.2 (20.5) |
| All values are means (SD). Rating range was 0-100. Negative and positive arousal values are ranging from -113 to 113 and are constrained to a maximum within-subject range of 141 for extreme values (typically much lower). | | | | | | | | | | |

| Table S2. Bayesian model comparison. Results showed that the dual learning rate model fitted the response data best in controls, whereas for MDD patients a simpler model with one learning rate performed better. Across all subjects, the simpler model provided the best model fit. | | | | | | |
| --- | --- | --- | --- | --- | --- | --- |
| Model | **MDD** | | **Controls** | | **All subjects** | |
|  | PP | XP | PP | XP | PP | XP |
| M1-rw1 | 2.5 | 0 | 2.3 | 0 | 1.4 | 0 |
| M2-rw1 | 2.5 | 0 | 2.3 | 0 | 1.4 | 0 |
| M3-rw1 | 57.7 | 99.6* | 25.9 | 2.4 | 55.4 | 98.6* |
| M4-rw1 | 2.5 | 0 | 2.4 | 0 | 1.4 | 0 |
| M5-rw1 | 3.8 | 0 | 4.1 | 0 | 2.4 | 0 |
| M1-rw2 | 2.5 | 0 | 2.3 | 0 | 1.4 | 0 |
| M2-rw2 | 2.5 | 0 | 2.3 | 0 | 1.4 | 0 |
| M3-rw2 | 19.6 | 0.4 | 51.6 | 97.6* | 31.4 | 1.4 |
| M4-rw2 | 2.5 | 0 | 2.4 | 0 | 1.4 | 0 |
| M5-rw2 | 3.8 | 0 | 4.4 | 0 | 2.4 | 0 |
| Asterisks indicate the winning model. rw1: Rescorla-Wagner with a single learning rate; rw2: Rescorla-Wagner with a dual learning rate; PP: expected posterior probability; XP: exceedance probability. | | | | | | |

| Table S3. Parameter prior means (variance) of the reinforcement learning model and the response models. | | | | | | |
| --- | --- | --- | --- | --- | --- | --- |
| Reinforcement learning model | **rw1** | | | **rw2** | | |
| $\boldsymbol{v}_{\boldsymbol{0}}$ (logit-space) | 0.66 (0.5) | | | 0.66 (0.5) | | |
| $\boldsymbol{\alpha}$ (logit-space) | 0.05 (1) | | | - | | |
| $\boldsymbol{\alpha}^{\boldsymbol{-}}$ (logit-space) | - | | | 0.05 (1) | | |
| $\boldsymbol{\alpha}^{\boldsymbol{+}}$ (logit-space) | - | | | 0.05 (1) | | |
| Response models | **M1** | **M2** | **M3** | | **M4** | **M5** |
| $\boldsymbol{\beta}_{\boldsymbol{0}}$ | 0 (4) | 0 (4) | 0 (4) | | 0 (4) | 0 (4) |
| $\boldsymbol{\beta}_{\boldsymbol{1}}$ | 0 (4) | 0 (4) | 0 (4) | | 0 (4) | 0 (4) |
| $\boldsymbol{\beta}_{\boldsymbol{2}}$ | 0 (4) | 0 (4) | 0 (4) | | 0 (4) | 0 (4) |
| $\boldsymbol{\beta}_{\boldsymbol{3}}$ | 0 (4) | 0 (4) | 0 (4) | | 0 (4) | 0 (4) |
| $\boldsymbol{\beta}_{\boldsymbol{4}}$ | 0 (4) | 0 (4) | 0 (4) | | 0 (4) | 0 (4) |
| $\boldsymbol{\beta}_{\boldsymbol{5}}$ | 0 (4) | - | - | | 0 (4) | - |
| $\boldsymbol{\beta}_{\boldsymbol{6}}$ | 0 (4) | - | - | | - | - |
| $\boldsymbol{\beta}_{\boldsymbol{7}}$ | 0 (4) | - | - | | - | - |
| $\boldsymbol{\xi}$ | log(3) (log(2)) | log(3) (log(2)) | log(3) (log(2)) | | log(3) (log(2)) | log(3) (log(2)) |

| Table S4. Averaged parameter estimates and parameter comparison between groups | | | | |
| --- | --- | --- | --- | --- |
|  | HC | MDD | Statistic | *p* value |
| $\boldsymbol{\alpha}$ | 0.052 (0.012) | 0.046 (0.009) | *t* (60) = 2.04^a^ | 0.046 |
| $\boldsymbol{\beta}_{\boldsymbol{0}}$ | -1.356 (0.156) | -1.362 (0.096) | *t* (61) = 0.184 | 0.854 |
| $\boldsymbol{\beta}_{\boldsymbol{1}}$ | -0.006 (0.030) | -0.023 (0.029) | *U* = 608^b^ | 0.122 |
| $\boldsymbol{\beta}_{\boldsymbol{2}}$ | 0.004 (0.026) | 0.015 (0.030) | *t* (61) = -1.490 | 0.141 |
| $\boldsymbol{\beta}_{\boldsymbol{3}}$ | 0.026 (0.050) | 0.005 (0.054) | *t* (61) = 1.638 | 0.106 |
| $\boldsymbol{\beta}_{\boldsymbol{4}}$ | -0.015 (0.074) | 0.001 (0.061) | *t* (61) = -0.903 | 0.370 |
| Mean (SD) within each group.  ^a^We removed one outlier in the MDD group, determined by visual inspection and a significant Grubb’s test  ^b^Mann-Whitney U-Tests were used for group comparison, because Shapiro-Wilk tests were significant in the MDD group. | | | | |

| **Table S5. Results of fMRI analyses: main effects of** $\boldsymbol{\varphi}_{\boldsymbol{t}}^{\boldsymbol{+}}$ **and** $\boldsymbol{\varphi}_{\boldsymbol{t}}^{\boldsymbol{-}}$ **during feedback processing (n = 63).** | | | | | | |
| --- | --- | --- | --- | --- | --- | --- |
|  | MNI coordinates [mm] | | | Significant activation | | Peak |
| Brain region | x | y | z | p_FWEc_ | k | Z |
| **Positive effect of** $\boldsymbol{\varphi}_{\boldsymbol{t}}^{\boldsymbol{+}}$ | | | | | | |
| Putamen_R | 21 | 10 | -10 | <.001 | 18272 | 6.52 |
| Frontal_Med_Orb_L | -7 | 50 | -12 |  |  | 6.28 |
| Caudate_R | 19 | 14 | 18 |  |  | 5.26 |
| Putamen_L | -15 | 18 | -4 | <.001 | 3072 | 6.35 |
| Frontal_Mid_2_L | -25 | 24 | 46 | <.001 | 1499 | 5.68 |
| Temporal_Inf_L | -47 | -6 | -26 | <.001 | 494 | 5.14 |
| Occipital_Mid_L | -39 | -74 | 36 | <.001 | 594 | 5.07 |
| Temporal_Inf_L | -57 | -46 | -16 | .03 | 120 | 4.42 |
| Temporal_Sup_L | -63 | -16 | 4 | .01 | 150 | 4.24 |
| **Negative effect of** $\boldsymbol{\varphi}_{\boldsymbol{t}}^{\boldsymbol{+}}$ | | | | | | |
| Frontal_Inf_Tri_R | 49 | 18 | 2 | <.001 | 1970 | 6.54 |
| Insula_R | 35 | 22 | -8 |  |  | 6.42 |
| Frontal_Inf_Orb_2_R | 45 | 44 | -8 |  |  | 6.42 |
| Frontal_Sup_Medial_R | 3 | 34 | 46 | <.001 | 1045 | 6.07 |
| Frontal_Sup_Medial_R | 3 | 40 | 40 |  |  | 4.91 |
| Cingulate_Mid_R | 3 | 34 | 46 |  |  | 4.85 |
| Insula_L | -29 | 26 | -10 | .001 | 215 | 4.77 |
| **Positive effect of** $\boldsymbol{\varphi}_{\boldsymbol{t}}^{\boldsymbol{-}}$ | | | | | | |
| Caudate_R | 21 | 8 | 16 | <.001 | 506 | 5.87 |
| Paracentral_Lobule_L | -13 | -28 | 60 | <.001 | 386 | 4.89 |
| Calcarine_R | 9 | -84 | 2 | <.001 | 367 | 4.65 |
| Caudate_L | -23 | -20 | 26 | <.001 | 283 | 4.50 |
| Temporal_Sup_R | 53 | -12 | 0 | <.001 | 265 | 4.49 |
| Occipital_Mid_L | -37 | -68 | 32 | .019 | 132 | 4.41 |
| Putamen_R | 25 | 14 | -2 | .016 | 137 | 4.35 |
| Fusiform_L | -33 | -40 | -20 | .013 | 142 | 4.20 |
| Occipital_Mid_L | -45 | -66 | 0 | .038 | 113 | 4.06 |
| Frontal_Sup_2_L | -15 | 42 | 48 | .001 | 231 | 3.96 |
| **Negative effect of** $\boldsymbol{\varphi}_{\boldsymbol{t}}^{\boldsymbol{-}}$ | | | | | | |
| Insula_R | 35 | 16 | -14 | <.001 | 425 | 5.35 |
| SupraMarginal_L | -69 | -36 | 32 | .011 | 148 | 4.49 |
| Insula_L | -29 | 20 | -18 | .002 | 205 | 4.41 |
| ACC_sup_R | 3 | 22 | 20 | <.001 | 446 | 4.25 |
| Significance level at whole-brain cluster-level pFWEc < 0.05, cluster-defining threshold pCDT < .001. Group effects are reported in Table 2.  Abbreviations: k, cluster size; R, right; L, left. | | | | | | |

| **Table S6. Correlation with CDI and anhedonia-subscale scores within patients (n=29).** | | | | | | |
| --- | --- | --- | --- | --- | --- | --- |
|  | MNI coordinates [mm] | | | Significant activation | | Peak |
| Brain region | x | y | z | p_FWEc_ | k | Z |
| **Negative correlation CDI with** $\boldsymbol{\varphi}_{\boldsymbol{t}}^{\boldsymbol{-}}$ | | | | | | |
| Insula_L | -45 | 0 | -4 | .027 | 107 | 4.80 |
| Postcentral_L | -67 | -20 | 24 | .023 | 111 | 4.45 |
| **Negative correlation between anhedonia CDI-subscale with** $\boldsymbol{\varphi}_{\boldsymbol{t}}^{\boldsymbol{-}}$ | | | | | | |
| Precentral_L | -27 | -14 | 54 | .012 | 127 | 4.77 |
| Insula_L | -37 | -10 | -2 | <.001 | 289 | 4.73 |
| SupraMarginal_R | 63 | -28 | 32 | .019 | 115 | 4.17 |
| Parietal_Inf_L | -51 | -30 | 52 | .015 | 104 | 3.90 |
| **Negative correlation CDI with** $\boldsymbol{\delta}_{\boldsymbol{t}}^{\boldsymbol{-}}$ | | | | | | |
| Fusiform_L | -35 | -44 | -22 | .001 | 230 | 4.75 |
| Temporal_Mid_L | -55 | -74 | 6 | .007 | 166 | 4.45 |
| Temporal_Inf_R | 49 | -58 | -6 | .045 | 112 | 4.39 |
| Fusiform_L | -35 | -10 | -32 | .006 | 169 | 4.27 |
| Parietal_Inf_R | 25 | -48 | 50 | .003 | 188 | 4.22 |
| **Negative correlation between anhedonia CDI-subscale with** $\boldsymbol{\delta}_{\boldsymbol{t}}^{\boldsymbol{-}}$ | | | | | | |
| Temporal_Inf_L | -37 | -12 | -34 | .001 | 214 | 5.60 |
| Postcentral_R | 29 | -32 | 36 | <.001 | 866 | 4.94 |
| Fusiform_R | 37 | -38 | -14 | .005 | 175 | 4.81 |
| Thal_MDm_R | 9 | -28 | 6 | <.001 | 268 | 4.54 |
| Postcentral_L | -33 | -36 | 56 | <.001 | 1492 | 4.51 |
| Temporal_Inf_R | 49 | -58 | -6 | <.001 | 272 | 4.43 |
| Temporal_Mid_L | -51 | -78 | 2 | <.001 | 389 | 4.31 |
| Cerebelum4_5_L | -23 | -38 | -22 | <.001 | 263 | 4.23 |
| Frontal_Sup_2_R | 31 | -6 | 66 | .001 | 238 | 4.02 |
| Cerebelum_6_R | 21 | -50 | -24 | .004 | 177 | 4.01 |
| **Positive correlation CDI with** $\boldsymbol{\varphi}_{\boldsymbol{t}}^{\boldsymbol{-}}$ | | | | | | |
| NS |  |  |  |  |  |  |
| **Positive correlation between anhedonia CDI-subscale with** $\boldsymbol{\varphi}_{\boldsymbol{t}}^{\boldsymbol{-}}$ | | | | | | |
| NS |  |  |  |  |  |  |
| **Positive correlation CDI with** $\boldsymbol{\delta}_{\boldsymbol{t}}^{\boldsymbol{-}}$ | | | | | | |
| NS |  |  |  |  |  |  |
| **Positive correlation between anhedonia CDI-subscale with** $\boldsymbol{\delta}_{\boldsymbol{t}}^{\boldsymbol{-}}$ | | | | | | |
| NS |  |  |  |  |  |  |
| Significance level at whole-brain cluster-level pFWEc < 0.05, cluster-defining threshold pCDT < .001.  Abbreviations: CDI, Child Depression Inventory; k, cluster size; R, right; L, left. | | | | | | |

| **Table S7. Results of fMRI analyses: main effects of magnitude-modulated prediction errors** $\boldsymbol{\delta}_{\boldsymbol{t}}^{\mathbf{+}}$ **and** $\boldsymbol{\delta}_{\boldsymbol{t}}^{\mathbf{-}}$ **during feedback processing (n = 63).** | | | | | | |
| --- | --- | --- | --- | --- | --- | --- |
|  | MNI coordinates [mm] | | | Significant activation | | Peak |
| Brain region | x | y | z | p_FWEc_ | k | Z |
| **Positive effect of** $\boldsymbol{\delta}_{\boldsymbol{t}}^{\boldsymbol{+}}$ | | | | | | |
| Ventr_Str_R | 9 | 10 | -14 | <.001 | 9071 | 6.67 |
| Frontal_Med_Orb_L | -5 | 48 | -14 |  |  | 6.65 |
| Frontal_Sup_2_L | -17 | 34 | 46 | <.001 | 1708 | 6.50 |
| Occipital_Inf_R | 27 | -90 | -2 | <.001 | 1392 | 5.92 |
| Postcentral_L | -17 | -30 | 60 | <.001 | 2783 | 5.46 |
| Occipital_Mid_L | -19 | -92 | 4 | <.001 | 320 | 4.92 |
| Temporal_Mid_L | -57 | -10 | -22 | <.001 | 260 | 4.79 |
| Temporal_Mid_R | 57 | -10 | -20 | .014 | 138 | 4.69 |
| Cingulate_Post_L | -1 | -46 | 32 | <.001 | 218 | 4.39 |
| Occipital_Mid_L | -43 | -74 | 32 | .013 | 140 | 4.32 |
| **Negative effect of** $\boldsymbol{\delta}_{\boldsymbol{t}}^{\boldsymbol{+}}$ | | | | | | |
| Frontal_Inf_Tri_R | 55 | 20 | 28 | <.001 | 1633 | 5.83 |
| Insula_R | 33 | 24 | 2 |  |  | 6.48 |
| Frontal_Inf_Tri_R | 55 | 20 | 28 | <.001 | 1633 | 5.83 |
| Frontal_Sup_Medial_L | 3 | 40 | 42 | <.001 | 424 | 4.76 |
| Insula_L | -29 | 22 | 8 | .001 | 228 | 4.31 |
| Frontal_Inf_Orb_2_R | 49 | 44 | -4 | .002 | 200 | 4.27 |
| **Positive effect of** $\boldsymbol{\delta}_{\boldsymbol{t}}^{\boldsymbol{-}}$ | | | | | | |
| Caudate_L | -19 | 2 | 20 | <.001 | 24740 | 7.18 |
| Caudate_R | 19 | -8 | 24 |  |  | 6.96 |
| Putamen_L | -13 | 12 | -14 | <.001 | 340 | 4.92 |
| Temporal_Mid_L | -57 | -32 | 6 | <.001 | 396 | 4.30 |
| Frontal_Sup_2_L | -19 | 26 | 52 | .005 | 189 | 4.20 |
| Frontal_Mid_2_L | -39 | 46 | -18 | .002 | 230 | 4.18 |
| **Negative effect of** $\boldsymbol{\delta}_{\boldsymbol{t}}^{\boldsymbol{-}}$ | | | | | | |
| Insula_L | -31 | 18 | -18 | <.001 | 1559 | >8 |
| Insula_R | 33 | 18 | -16 | <.001 | 2297 | 7.63 |
| Frontal_Sup_Medial_L | 7 | 32 | 26 | <.001 | 4963 | 6.64 |
| Red_N_L | -3 | -24 | 26 | <.001 | 538 | 6.59 |
| SupraMarginal_R | 59 | -46 | 46 | <.001 | 761 | 5.60 |
| SupraMarginal_L | -65 | -46 | 38 | <.001 | 380 | 5.29 |
| Temporal_Mid_R | 53 | -32 | -6 | <.001 | 343 | 5.17 |
| Frontal_Sup_2_R | 19 | 54 | 24 | .013 | 157 | 3.99 |
| Significance level at whole-brain cluster-level pFWEc < 0.05, cluster-defining threshold pCDT < .001. Abbreviations: k, cluster size; R, right; L, left. | | | | | | |

| **Table S8. Task activation across groups (n=60).** | | | | | | |
| --- | --- | --- | --- | --- | --- | --- |
|  | MNI coordinates [mm] | | | Significant activation | | Peak |
| Brain region | x | y | z | p_FWE_ | k | Z |
| **Hit-miss** | | | | | | |
| Caudate_R | 19 | 8 | 20 | < .001 | 2752 | 7.75 |
| Caudate_L | -19 | -10 | 24 | < .001 | 714 | 7.66 |
| Putamen_L | -21 | 12 | -8 | < .001 | 791 | 7.15 |
| Paracentral_Lobule_L | -7 | -36 | 62 | < .001 | 1951 | 6.96 |
| Cuneus_L | -23 | -52 | 20 | < .001 | 167 | 6.63 |
| Calcarine_L | -21 | -86 | 6 | < .001 | 348 | 6.59 |
| Hippocampus_R | 43 | -22 | -18 | < .001 | 82 | 6.43 |
| Frontal_Sup_2_L | -21 | 32 | 46 | < .001 | 369 | 6.31 |
| Frontal_Med_Orb_L | -7 | 46 | -16 | < .001 | 250 | 6.24 |
| Temporal_Sup_R | 67 | -8 | -2 | < .001 | 186 | 5.95 |
| Precuneus_L | -11 | -54 | 8 | < .001 | 123 | 5.90 |
| Calcarine_R | 15 | -12 | 46 | < .001 | 51 | 5.72 |
| **Miss-hit** | | | | | | |
| Frontal_Sup_Media_R | 5 | 28 | 54 | < .001 | 784 | 7.49 |
| Frontal_Sup_Media_R | 7 | 36 | 30 | < .001 |  | 6.87 |
| Insula_R | 35 | 18 | -18 | < .001 | 1195 | 7.43 |
| Insula_L | -31 | 20 | -14 | < .001 | 579 | 7.38 |
| Temporal_Mid_R | 57 | -30 | -8 | < .001 | 56 | 6.21 |
| SupraMarginal_R | 63 | -48 | 36 | < .001 | 67 | 5.40 |
| Significant clusters on whole-brain level in the second-level analyses for the contrasts reward-miss and miss-reward. This analysis aimed at revealing the task-relevant network across groups and guide the selection of ROIs for further analysis, thus we included the CDI score as covariate.  Significance level at whole-brain voxel-level p_FWE_ < 0.05, k > 50.  Abbreviations: CDI, Child Depression Inventory; k, cluster size; R, right; L, left. | | | | | | |

| **Table S9. Expected value (Q)-signaling across both groups.** | | | | | | |
| --- | --- | --- | --- | --- | --- | --- |
|  | MNI coordinates [mm] | | | Significant activation | | Peak |
| Brain region | x | y | z | p_FWEc_ | k | Z |
| **Positive effect of Q+** | | | | | | |
| Insula_R | 39 | 28 | -4 | < .001 | 21473 | 6.07 |
| Raphe_D | -5 | -30 | -14 |  |  | 5.87 |
| Vermis_6 | 7 | -66 | -10 |  |  | 5.66 |
| Insula_R | 29 | 18 | -16 |  |  | 5.24 |
| Unknown | 7 | -30 | -14 |  |  | 5.21 |
| Pallidum_R | 11 | 4 | -4 |  |  | 5.21 |
| Thal_PuM_R | 3 | -24 | 10 |  |  | 5.20 |
| ACC_pre_L | 1 | 42 | 20 |  |  | 5.02 |
| SupraMarginal_R | 53 | -42 | 30 | .003 | 198 | 3.90 |
| Postcentral_L | -49 | -14 | 50 | .005 | 183 | 3.90 |
| Precentral_R | 39 | -10 | 48 | .002 | 209 | 3.86 |
| **Positive effect of Q-** | | | | | | |
| NS |  |  |  |  |  |  |
| Significance level at whole-brain cluster-level p_FWEc_ < 0.05, cluster-defining threshold p_CDT_ < .001. Abbreviations:k, cluster size; R, right; L, left. | | | | | | |

| **Table S10. Positive effect of all feedback events.** | | | | | | |
| --- | --- | --- | --- | --- | --- | --- |
|  | MNI coordinates [mm] | | | Significant activation | | Peak |
| Brain region | x | y | z | p_FWE(c)_ | k | Z |
| Occipital_Inf_R | 29 | -80 | -16 | < .001 | 26989 | > 8 |
| Frontal_Sup_Medial_R | 5 | 28 | 44 | < .001 | 1931 | > 8 |
| Hippocampus_L | -25 | -28 | -6 | < .001 | 139 | 7.47 |
| Frontal_Inf_Orb_2_L | -37 | 20 | -6 | < .001 | 402 | 7.36 |
| Parietal_Inf_L | -49 | -44 | 54 | < .001 | 1553 | 7.14 |
| Vermis_4_5 | 1 | -36 | -2 | < .001 | 142 | 6.73 |
| Caudate_R | 9 | 24 | 0 | < .001 | 145 | 6.66 |
| Frontal_Mid_2_L | -39 | 60 | -4 | < .001 | 1071 | 6.43 |
| Cerebellum_10_R | 23 | -40 | -44 | < .001 | 115 | 6.34 |
| Postcentral_L | -65 | -10 | 30 | < .001 | 133 | 5.91 |
| This contrast was used to identify activity in the visual areas for the DCM analysis.  Significance level at whole-brain whole-brain p_FWE_ < 0.05, minimum cluster size k > 100. Abbreviations:DCM, dynamic causal modeling; k, cluster size; R, right; L, left. | | | | | | |

### Supplementary References

1.	Knutson B, Katovich K, Suri G (2014): Inferring affect from fMRI data. *Trends in cognitive sciences*. 18:422-428.

2.	Rescorla RA, Wagner AR (1972): A theory of Pavlovian conditioning: Variations in the effectiveness of reinforcement and nonreinforcement. *Classical conditioning II: Current research and theory*. 2:64-99.

3.	O'Doherty JP, Dayan P, Friston K, Critchley H, Dolan RJ (2003): Temporal difference models and reward-related learning in the human brain. *Neuron*. 38:329-337.

4.	McClure SM, Berns GS, Montague PR (2003): Temporal prediction errors in a passive learning task activate human striatum. *Neuron*. 38:339-346.

5.	Palminteri S, Justo D, Jauffret C, Pavlicek B, Dauta A, Delmaire C, et al. (2012): Critical roles for anterior insula and dorsal striatum in punishment-based avoidance learning. *Neuron*. 76:998-1009.

6.	Rigoli F, Chew B, Dayan P, Dolan RJ (2016): Multiple value signals in dopaminergic midbrain and their role in avoidance contexts. *Neuroimage*. 135:197-203.

7.	Bayer HM, Glimcher PW (2005): Midbrain dopamine neurons encode a quantitative reward prediction error signal. *Neuron*. 47:129-141.

8.	Cao Z, Bennett M, Orr C, Icke I, Banaschewski T, Barker GJ, et al. (2019): Mapping adolescent reward anticipation, receipt, and prediction error during the monetary incentive delay task. *Human brain mapping*. 40:262-283.

9.	Beierholm U, Guitart-Masip M, Economides M, Chowdhury R, Düzel E, Dolan R, et al. (2013): Dopamine modulates reward-related vigor. *Neuropsychopharmacology*. 38:1495.

10.	Dudman JT, Krakauer JW (2016): The basal ganglia: from motor commands to the control of vigor. *Current opinion in neurobiology*. 37:158-166.

11.	Bestmann S, Ruge D, Rothwell J, Galea JM (2014): The role of dopamine in motor flexibility. *Journal of cognitive neuroscience*. 27:365-376.

12.	Niv Y, Daw ND, Joel D, Dayan P (2007): Tonic dopamine: opportunity costs and the control of response vigor. *Psychopharmacology*. 191:507-520.

13.	Lawson RP, Seymour B, Loh E, Lutti A, Dolan RJ, Dayan P, et al. (2014): The habenula encodes negative motivational value associated with primary punishment in humans. *Proceedings of the National Academy of Sciences*. 111:11858-11863.

14.	Bunzeck N, Düzel E (2006): Absolute coding of stimulus novelty in the human substantia nigra/VTA. *Neuron*. 51:369-379.

15.	Stephan KE, Penny WD, Daunizeau J, Moran RJ, Friston KJ (2009): Bayesian model selection for group studies. *Neuroimage*. 46:1004-1017.

16.	Power JD, Barnes KA, Snyder AZ, Schlaggar BL, Petersen SE (2012): Spurious but systematic correlations in functional connectivity MRI networks arise from subject motion. *Neuroimage*. 59:2142-2154.
